# Prediction of Covid-19 Infections Through December 2020 for 10 US States Incorporating Outdoor Temperature and School Re-Opening Effects-August Update

**DOI:** 10.1101/2020.09.14.20193821

**Authors:** Ty Newell

**Affiliations:** Emeritus Professor of Mechanical Engineering, Department of Mechanical Science and Engineering, University of Illinois, Urbana Illinois USA

## Abstract

End-of-August updates for Covid-19 infection case predictions for 10 US States (NY, WA, GA, IL, MN, FL, OH, MI, CA, and NC) are compared to actual data. Several states that experienced significant summer surges gained control of accelerating infection spread during August. The US as a whole and the 10 States investigated continue to follow periods of linear infection growth that defines a boundary separating accelerated infection growth and infection decay.

August 31 predictions (initiated July 27, 2020) for four States (NY, WA, MI and MN) are within 10% of actual data. Predictions for four other States (GA, IL, CA, and OH) agree between 10 and 20% of actual data. Predictions for two States (FL and NC) are greater than 20% different from actual data. Systematic differences between predictions and actual data are related to the impact of the June-July summer surge, and human behavior reactions (ie, increased face mask usage and distancing) to accelerated infection growth.

Outdoor temperature effects and school re-opening effects are not apparent nor expected for August. Human behavior parameters (Social Distance Index, SDI, and disease transmission efficiency, G, parameters) are adjusted to mirror August data. Comparisons of actual versus predicted daily new infection cases display the complexity of SARS-CoV-2 transmission.

## Introduction

August displays a tendency of the United States to follow a push-pull path across a linear infection growth boundary that is a demarcation between accelerating growth and decay of Covid-19 infections. The push-pull across the linear infection growth boundary is a result of human reactions to news of strong infection growth causing increased social distancing and increased inter-personal protection (eg, face masks, 6ft distancing, increased ventilation and filtration). Countering infection protection measures is a desire to return to “normal” human interactions with relaxed disease transmission control behaviors.

Infection trends for 10 US States (NY, WA, GA, IL, MN, FL, OH, MI, CA, NC) are compared to infection predictions described in a previous report (1) that initiated infection predictions as of July 27, 2020. The infection prediction model includes the effects of an anticipated outdoor temperature impact and school re-openings.

An outdoor temperature effect was observed during the spring of 2020 that indicated reduced SARS-CoV-2 transmission efficiency when outdoor temperatures were between 50F (10C) and 70F (21C) (2). Possible reasons for the observed outdoor temperature effect are increased outdoor activities, increased building ventilation (eg, window openings), and decreased building occupancy due to increased outdoor activities. With the exception of Washington, which has been in the 50F to 70F temperature window during August, all other States modeled have had average ambient temperatures above 70F. Some of these States will drop into the temperature window during September.

Model predictions for four States (NY, WA, MN, MI) agreed within 10% of actual data at the end of August. Four States (GA, IL, CA, and OH) predictions are between 10 and 20% of actual data at the end of August. Two States (FL and NC) predictions differed by more than 20% from actual data. The differences between prediction and actual data are systematic and related to human behavior as States reacted to the June-July summer infection surge. States that were minimally impacted by the summer surge (NY, WA, MN, MI) moved more closely along the linear infection growth boundary. States that were more strongly impacted by the surge instituted strong control measures (eg, face mask and distancing regulations) that moved States from the infection acceleration side of the linear infection growth boundary to the infection decay region during August.

Infection Parameter (IP) trends during August are discussed for the US and 10 States. IP is a function of two human behaviors that are characterized by a Social Distance Index (SDI) and a disease transmission efficiency (G). The SDI is an independently obtained parameter based on anonymous cell phone and vehicular GPS data, and characterizes how much people travel outside their home. The disease transmission efficiency describes inter-personal conditions (eg, wearing face masks, maintaining 6ft spacing, increased fresh air ventilation and air filtration, etc).

Systematic differences between model predictions for the end of August and actual data reflect benefits achieved by implementing disease transmission controls. That is, one can interpret the difference between predicted human behavior and actual as showing the number of August infections that were avoided. Adjustments to the prediction model to reflect actual August IP trends are made, without any adjustments to model parameters beyond the end of August. As discussed in the initial report (1), physical school re-openings in September may increase infection growth as increased social interaction occurs. September is also a month in which several of the more northern States examined will enter a beneficial outdoor temperature window between 50F and 70F that may help reduce infection growth, in contrast to the 1918 pandemic’s fall infection surge.

Finally, new daily infection predictions and actual data for each State are compared. New daily infections display the complexity and variety of infection transmission among the 10 States modeled. Overall, Covid-19’s spread is strongly impacted by human behavior as characterized by the prediction model’s gross human interaction (SDI) and interpersonal interaction (G). Without strong guidance and mandates, a populace is likely to alternately move across the linear infection growth boundary as observed in the US and several other countries that lack a coherent and coordinated infection control plan.

## Background – Linear Infection Growth Models

Linear growth of SARS-CoV-2 infections is observed in many countries. Other investigators (6, 7, 8) have noticed linear infection growth and noted that traditional disease transmission models do not naturally display linear infection growth trends.

Ansumali and Prakash (6) observed “plateauing” rather than peaking of new daily infection cases resulting in linear growth of cumulative infections. The investigators incorporated parameters into an SEIR model which resulted in linear infection growth. The linear growth deviation from expected exponential growth characteristics of SEIR models is attributed to a persistence parameter that characterizes continuing but reduced disease transmission pathways. The authors also noted the importance of the 7 day incubation period in contrast to much shorter incubations for influenza and SARS.

Thurner, Klimek and Hanel (7) use a network model to examine how exponential disease growth can transition to a linear infection growth path. The authors’ discrete, network modeling approach provides a formulation basis that is flexible for exploring disease transmission modes in a mechanistic manner. The authors provide an explanation for the transition from exponential infection growth to linear infection growth through a contact parameter, similar to the social distance index used in the present model.

Grinfeld and Mulheran (8) formulate a two equation model that displays linear infection growth characteristics. The authors describe a process in which daily infection information relayed to a populace directly impacts their social interactions. The authors describe linear growth as a characteristic of countries without significant government intervention and use Sweden’s early “herd immunity” experiment as an example of linear growth.

The single equation model formulated for the present investigation is described in detailed in the appendices of reference (2). The model assumes infection growth rate is proportional to the number of infectious cases within a given populace. The proportionality factor is based on an Infection Parameter (IP) that incorporates Covid-19’s incubation period (assumed as 7 days) and infectious period (assumed as 14 days). IP is related to two human behavior parameters, a Social Distance Index (SDI) and a disease transmission efficiency parameter (G). The SDI reflects gross movement of people while G accounts for inter-personal behaviors (eg handshaking, hugging, face mask wearing, fresh air ventilation levels, etc). Increasing SDI and decreasing G move IP to its lower limit of 1 (zero infection growth rate).

The present model directly predicts the existence of linear infection growth. Additionally, linear infection growth is shown to be a boundary separating regions of accelerating and decreasing infections. The linear boundary is defined by an Infection Parameter (IP) value of “e” (~2.72), which conceptually represents the ratio of infections resulting from an infectious person over their 14 day infectious period.

The linear infection growth boundary is not defined by its slope. That is, the linear boundary is independent of the amount of daily new infections. Human reaction to news of accelerated infection growth, however, is related to the number of new infection cases. Strong human reaction to uncontrolled infection growth results in strong human countermeasures that can move a populace further into the decaying infection region.

All data used for modeling and correlations are publicly available. Data for US total infections are obtained from Worldometers.com that compiles data from multiple sources (3). Data for US States total infections are obtained from 91-divoc.com, a website formulated by a University of Illinois faculty member that utilizes data from Johns Hopkins University (4). Data for the Social Distance Index is obtained from the University of Maryland’s Transportation Institute website (5).

## Infection Parameter (IP), Social Distance Index (SDI) and Disease Transmission Efficiency (G)

Figure 1 shows total US Covid-19 disease cases. Three regions of linear growth are identified on the plot, including an initial linear growth period during May, a second linear period in July, and a more recent linear period in August. The initial linear infection period occurred as early infection growth States (eg, NY, WA, IL, MI) were able to gain control and stabilize infection transmission rates. The second linear period in late July occurred after the “summer surge” in the latter half of June and early July. Continued implementation of disease transmission control measures in August further reduced the slope of infection growth to the third (current) linear infection growth period.

**Figure 1.**
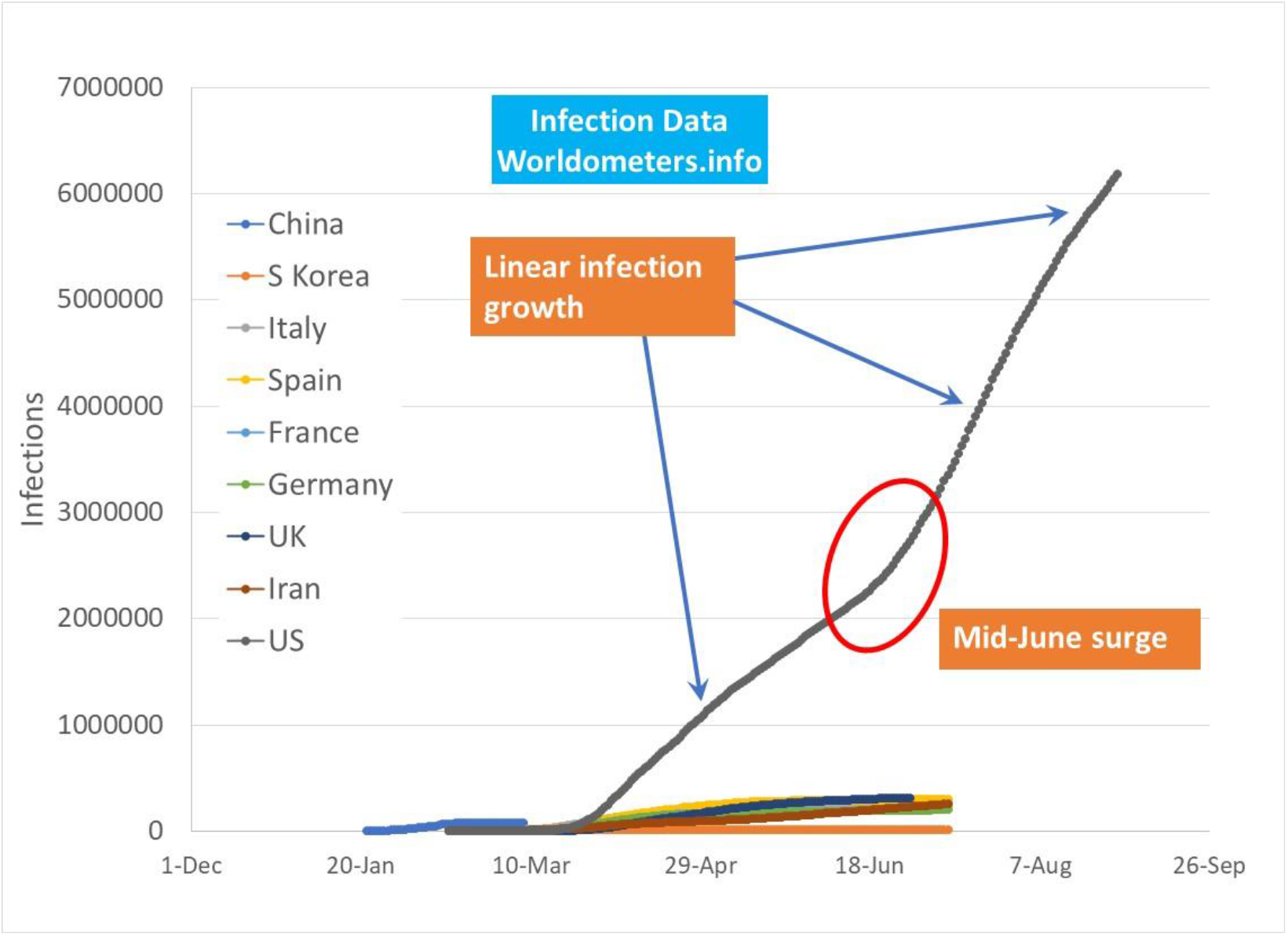
*US total infections as of July 31 2020 with linear infection path regions before and after mid-June “summer surge”*.

The present infection growth model directly predicts linear infection growth with an Infection Parameter (IP) value of “e” (~2.72). Figure 2 shows IP for the US since March 2020. Initially, IP was very high prior to the US populace implementing isolation and personal protection measures. During April and May, the US moved along the linear infection growth boundary of 2.72. An infection growth surge occurred in mid-June, reflected in an increase of IP above 2.72. The summer surge was a combination of increased outdoor temperatures in more northern States that increased indoor occupancy with elevated airborne disease transmission, and premature, poorly managed business re-openings in southern States (2).

**Figure 2.**
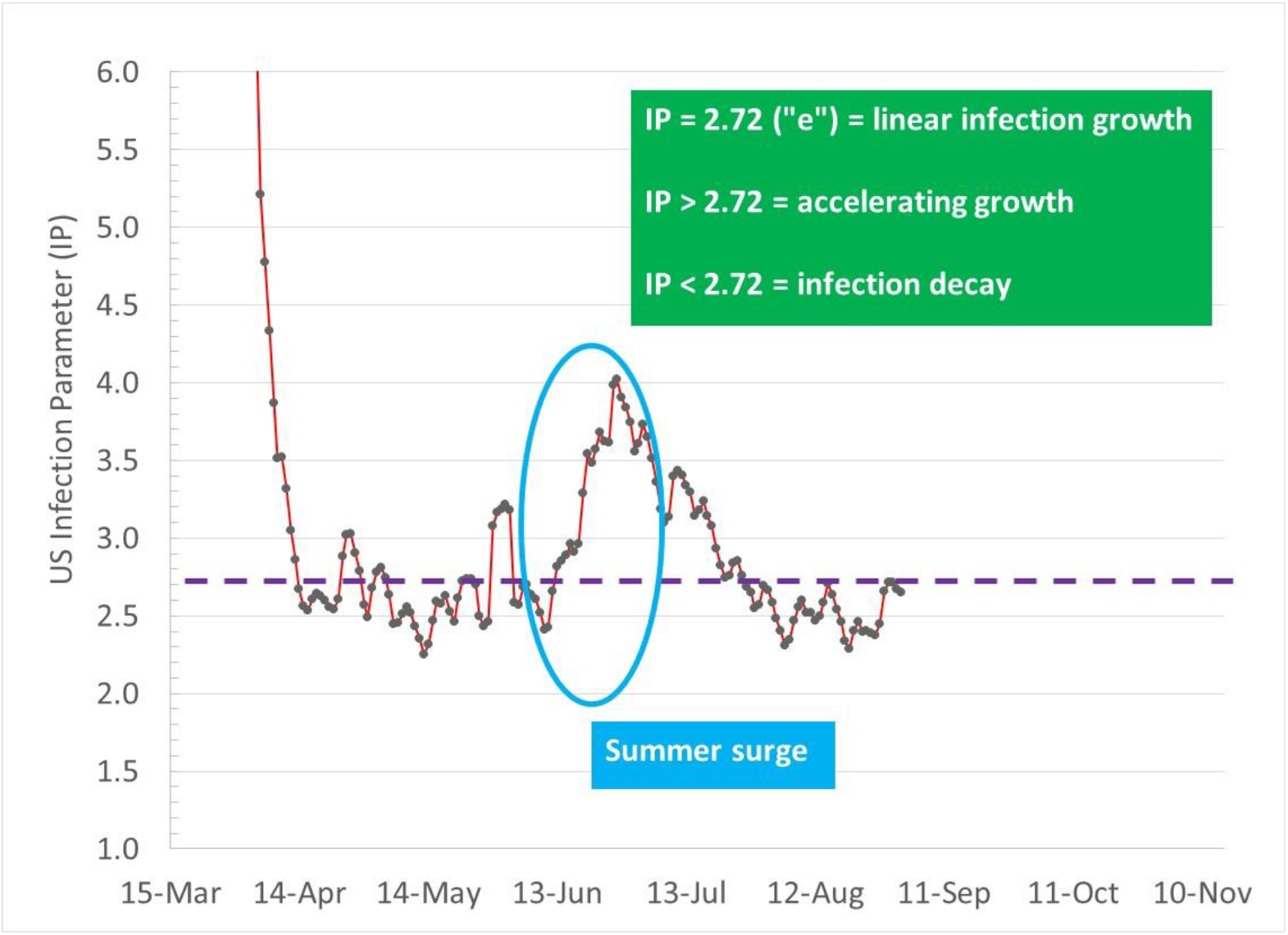
**US Infection Parameter (IP) showing tendency to move along linear total infection growth boundary (IP=2.72). The mid-June “summer surge” caused by a combination of outdoor temperature increase above 70F and pre-mature business openings is followed by local populace actions to reduce infection growth back to the linear growth boundary*.*

Figure 3 shows IP trends for the 10 States investigated in this report. State trends follow US IP trends, however, individual State trends are varied. Florida, for example, suffered severe infection growth during the June-July surge that resulted in business closures, strict face mask regulations, and an overall reaction that moved Florida to sustained IP levels below the 2.72 limit in August that reduced infections well below predicted infections.

**Figure 3.**
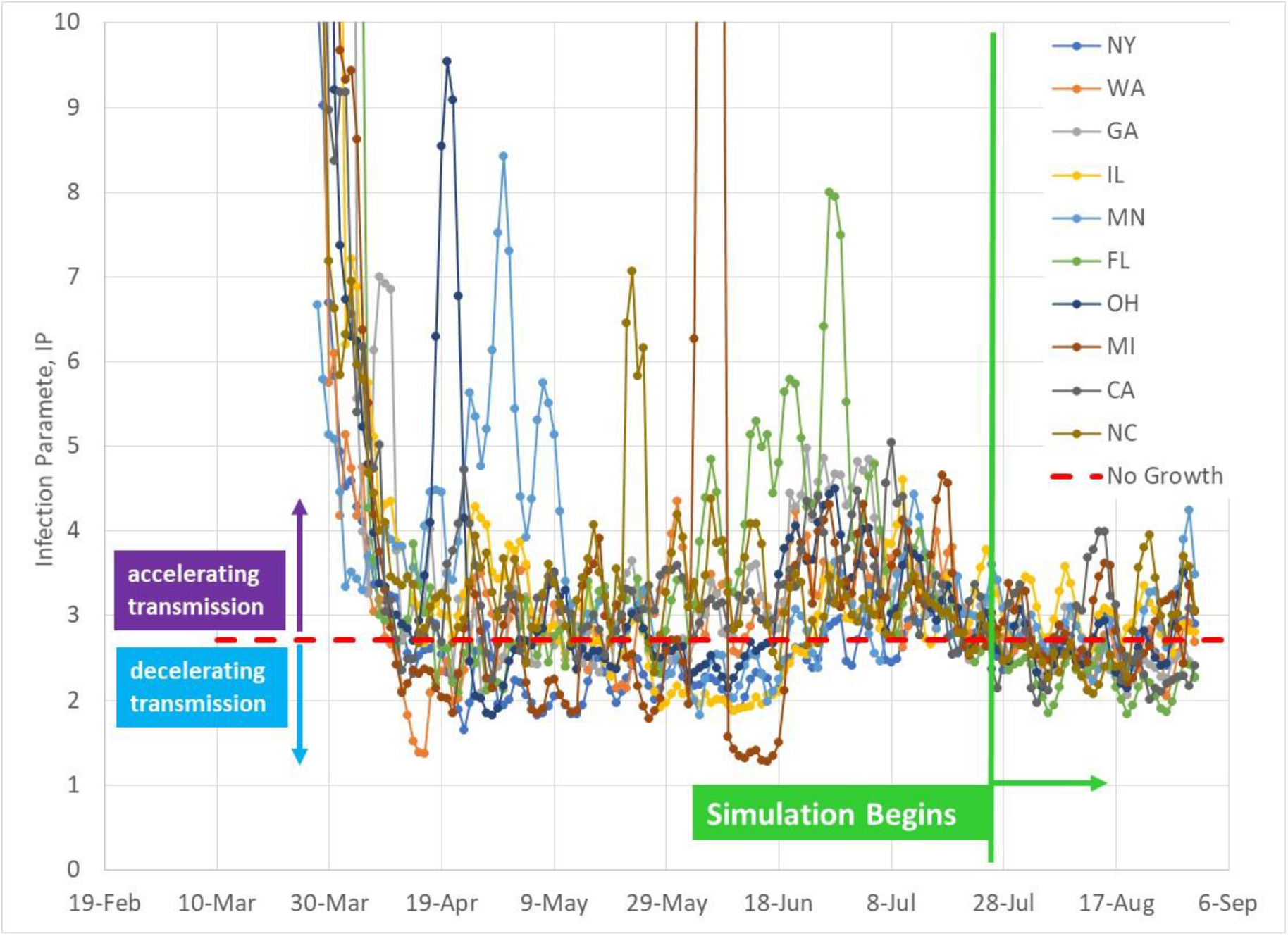
*Infection Parameter for 10 US States through August 31, 2020*.

Figures 4 and 5 display how IP is a function of human behavior through two independent parameters, SDI (Social Distance Index) and G (disease transmission efficiency). Figure 4 is an expanded view of IP versus SDI and G. Beginning in March 2020, IP levels were high prior to isolation and introduction of inter-personal protection measures (no handshaking, no hugging, face masks, etc). The present model assumes this initial trend (open blue circle data points) to define a disease transmission efficiency parameter (G) value of 1 based on average US IP data. As the US reached its maximum isolation level (maximum SDI) in April, inter-personal protection guidance measures such as 6 ft distancing, wearing face masks, and reduced indoor occupant densities were being implemented. The result of inter-personal protection measures is a decrease of the disease transmission efficiency parameter.

**Figure 4.**
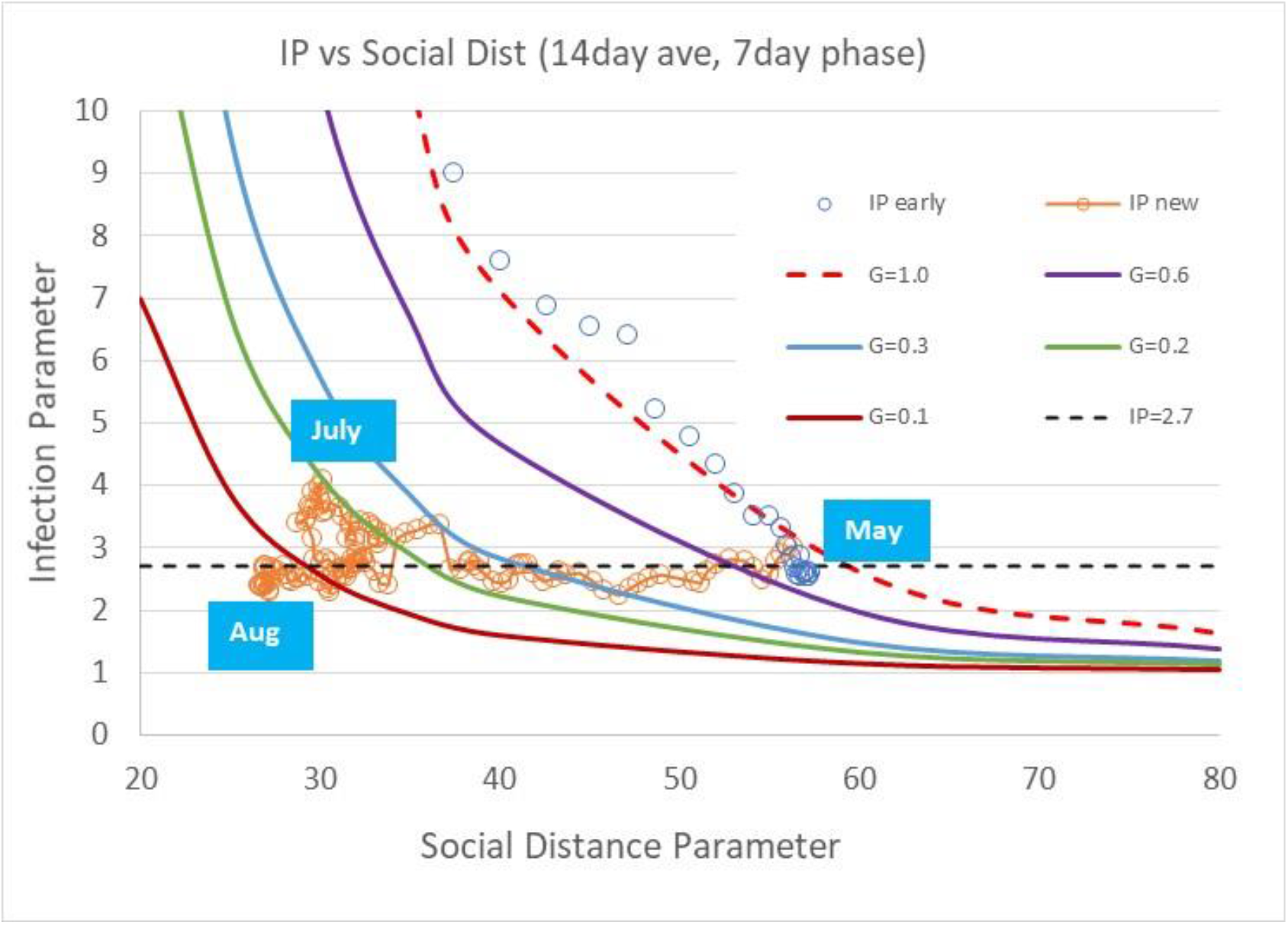
*US average Infection Parameter as a function of Social Distance Index (SDI) and diseases transmission efficiency parameter (G)*

**Figure 5.**
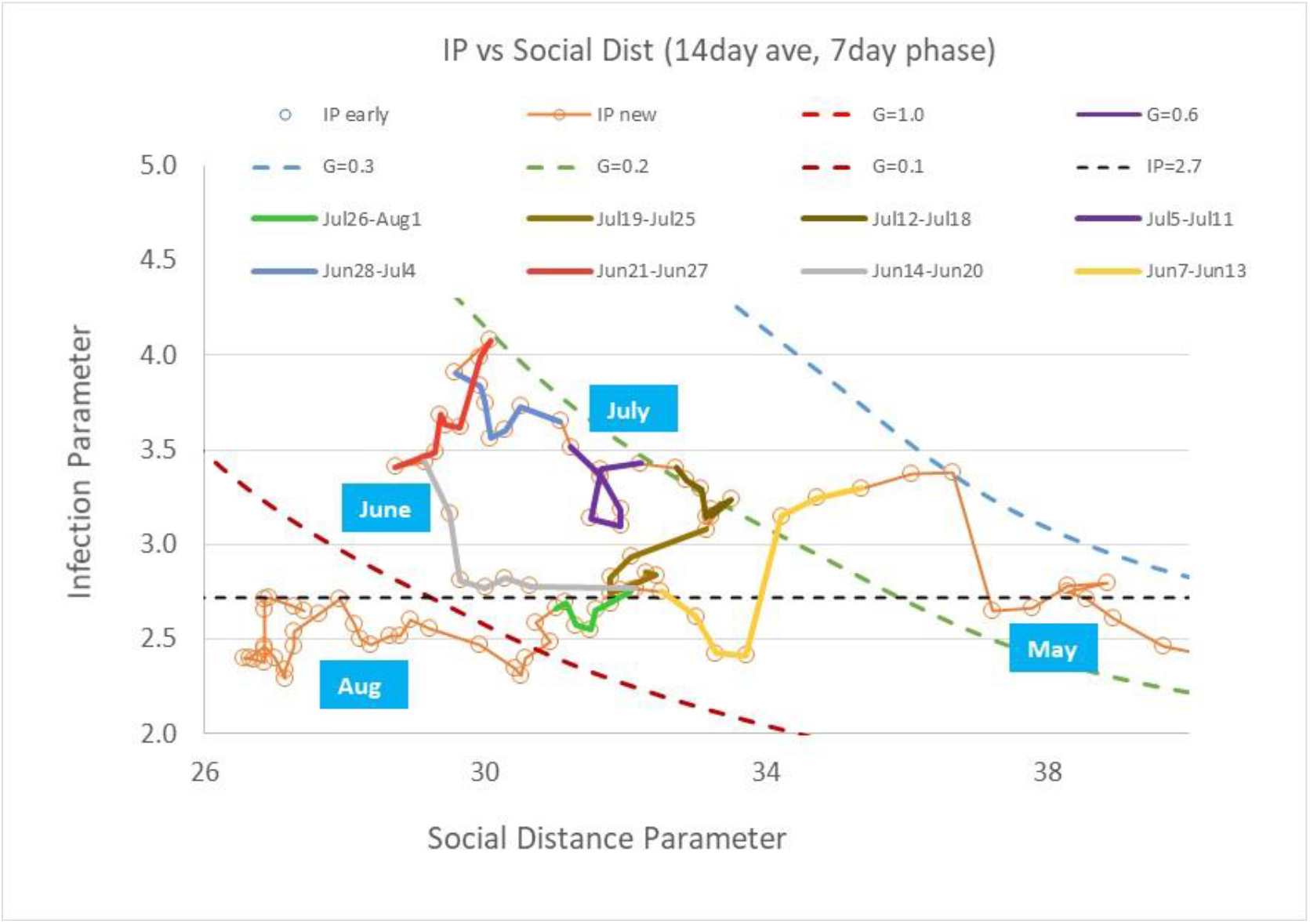
*US average Infection Parameter as a function of Social Distance Index (SDI) and disease transmission efficiency parameter (G) from May through August 2020*

Figure 4 shows the observed tendency of a populace (the US as a whole, several US States, and several other countries) that have not implemented stringent control measures to follow along the linear infection path boundary with an IP of 2.72. Figure 5 shows an enlarged view of IP versus SDI and G trends for May through August 2020. As the US re-opened business activities, SDI was reduced along an IP value of 2.72. By mid-June 2020, the occurrence of warm weather forced people indoors to air conditioning (which was not available to people during the 1918 Pandemic) where the disease transmission efficiency sharply increased as observed by the latter two week IP trends in June in Figure 5. Simultaneously, southern States experienced significant increases in IP due to continued reduction of SDI (ie, increased social interactions) without an accompanying reduction in disease transmission efficiency, G.

The US reaction to the infection surge in June and July was an increase of SDI (increased isolation) at constant transmission efficiency, G, followed by renewed transmission control efforts (reduced indoor activities, increased face mask regulations and increased participation of people in control measures). By the end of July, when the current model predictions were initiated, the US had reduced IP slightly below the linear limit of 2.72. Since the beginning of August, IP levels below 2.72 have been sustained, with a reduction of new daily infection cases. Social interactions have increased through August (decreased SDI) coupled with more stringently followed inter-personal protection measures (reduced G).

As of the end of August, the US has stabilized near the 2.72 IP linear boundary as shown in Figure 5. Movement of IP in September should begin revealing whether increased physical school and business re-opening attempts coupled with outdoor temperature changes significantly impact Covid-19 infections.

## Comparison of Predicted and Actual August Infection Cases

Table 1 and Figures 6-9 compare total infection cases for 10 US States as of August 31, 2020. The prediction model’s initial prediction started on July 27, 2020. Four prediction cases are shown in Table 1 and Figures 6-9:

1. No school re-openings; fall season temperature effect
2. No school re-openings; no fall season temperature effect
3. School re-openings; fall season temperature effect
4. School re-openings; no fall season temperature effect

**Table 1.** Total Infections and New Daily Infections as of July 27, 2020 August 31, 2020 actual and August 31, 2020 predictions for 10 US States.

| August 31, 2020 |  |  |  |  |  |  |  |  |  |  |
| --- | --- | --- | --- | --- | --- | --- | --- | --- | --- | --- |
| Total Infections | NY | WA | GA | IL | MN | FL | OH | MI | CA | NC |
| July 27 2020 | 411736 | 52635 | 167953 | 172663 | 51153 | 423855 | 84073 | 86661 | 452288 | 112937 |
| No School T Effect Actual | 434756 | 74635 | 270471 | 236724 | 75864 | 623471 | 123155 | 113025 | 712475 | 167309 |
| No School T Effect Predict | 436527 | 72021 | 315958 | 273980 | 82699 | 920599 | 148236 | 122356 | 803355 | 207747 |
| No School T Effect %Diff | -0.4% | 3.5% | -16.8% | -15.7% | -9.0% | -47.7% | -20.4% | -8.3% | -12.8% | -24.2% |
| No School No T Actual | 434756 | 74635 | 270471 | 236724 | 75864 | 623471 | 123155 | 113025 | 712475 | 167309 |
| No School No T Predict | 436527 | 85301 | 315958 | 275891 | 83174 | 920599 | 149484 | 123060 | 954929 | 207747 |
| No School No T %Diff | -0.4% | -14.3% | -16.8% | -16.5% | -9.6% | -47.7% | -21.4% | -8.9% | -34.0% | -24.2% |
| School T Effect Actual | 434756 | 74635 | 270471 | 236724 | 75864 | 623471 | 123155 | 113025 | 712475 | 167309 |
| School T Effect Predict | 436527 | 72021 | 315958 | 273980 | 82699 | 920599 | 148236 | 122356 | 803355 | 207747 |
| School T Effect %Diff | -0.4% | 3.5% | -16.8% | -15.7% | -9.0% | -47.7% | -20.4% | -8.3% | -12.8% | -24.2% |
| School No T Effect Actual | 434756 | 74635 | 270471 | 236724 | 75864 | 623471 | 123155 | 113025 | 712475 | 167309 |
| School No T Effect Predict | 436527 | 85301 | 315958 | 275891 | 83174 | 920599 | 149484 | 123060 | 954929 | 207747 |
| School No T Effect %Diff | -0.4% | -14.3% | -16.8% | -16.5% | -9.6% | -47.7% | -21.4% | -8.9% | -34.0% | -24.2% |
| New Daily Infections |  |  |  |  |  |  |  |  |  |  |
| July 27 2020 | 536 | 786 | 2765 | 1541 | 862 | 9344 | 889 | 1039 | 5836 | 1516 |
| No School T Effect Actual | 659 | 466 | 2031 | 1931 | 795 | 2949 | 1072 | 763 | 5273 | 1559 |
| No School T Effect Predict | 681 | 314 | 3660 | 1873 | 698 | 12837 | 1233 | 699 | 8433 | 2452 |
| No School T Effect %Diff | -3.4% | 32.6% | -80.2% | 3.0% | 12.2% | -335.3% | -15.0% | 8.4% | -59.9% | -57.3% |
| No School No T Actual | 659 | 466 | 2031 | 1931 | 795 | 2949 | 1072 | 763 | 5273 | 1559 |
| No School No T Predict | 681 | 848 | 3660 | 2540 | 866 | 12837 | 1670 | 945 | 13011 | 2452 |
| No School No T %Diff | -3.4% | -81.8% | -80.2% | -31.5% | -8.9% | -335.3% | -55.8% | -23.9% | -146.7% | -57.3% |
| School T Effect Actual | 659 | 466 | 2031 | 1931 | 795 | 2949 | 1072 | 763 | 5273 | 1559 |
| School T Effect Predict | 681 | 314 | 3660 | 1873 | 698 | 12837 | 1233 | 699 | 8433 | 2452 |
| School T Effect %Diff | -3.4% | 32.6% | -80.2% | 3.0% | 12.2% | -335.3% | -15.0% | 8.4% | -59.9% | -57.3% |
| School No T Effect Actual | 659 | 466 | 2031 | 1931 | 795 | 2949 | 1072 | 763 | 5273 | 1559 |
| School No T Effect Predict | 681 | 848 | 3660 | 2540 | 866 | 12837 | 1670 | 945 | 13011 | 2452 |
| School No T Effect %Diff | -3.4% | -81.8% | -80.2% | -31.5% | -8.9% | -335.3% | -55.8% | -23.9% | -146.7% | -57.3% |

**Figure 6.**
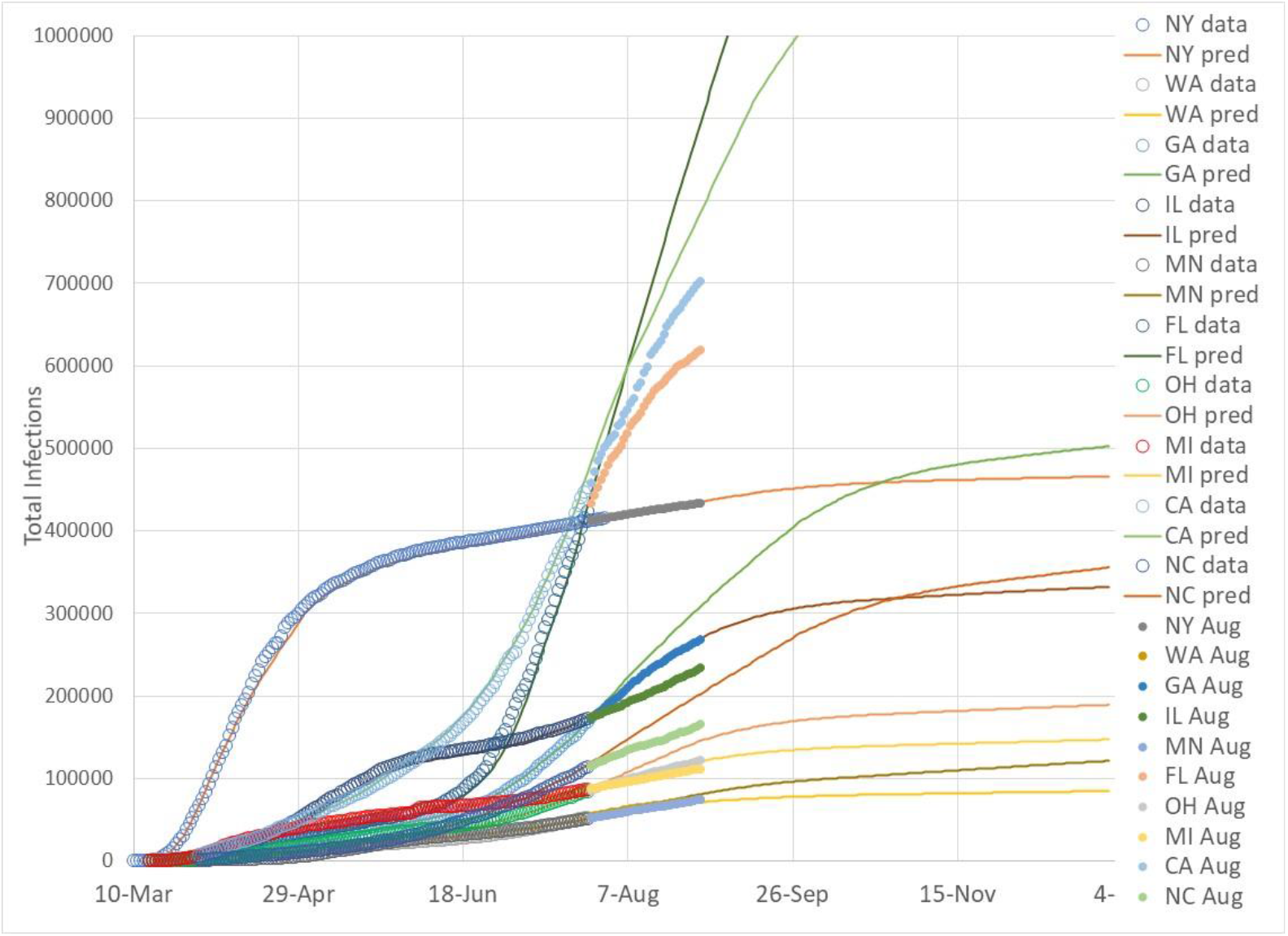
*Comparison of prediction and actual August total infection data for 10 State for No School with Temperature effect*.

**Figure 7.**
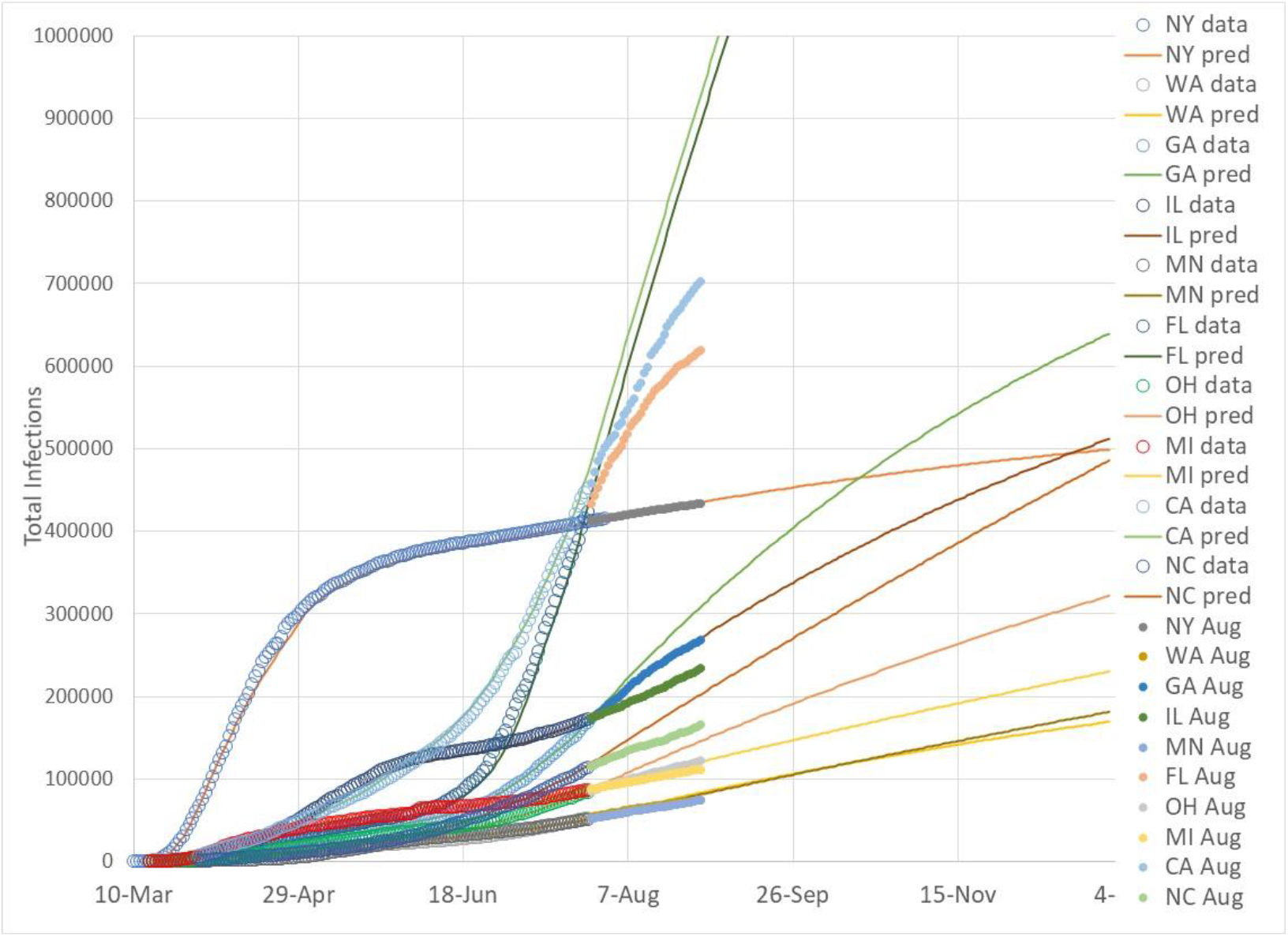
*Comparison of prediction and actual August total infection data for 10 State for No School with No Temperature effect*.

**Figure 8.**
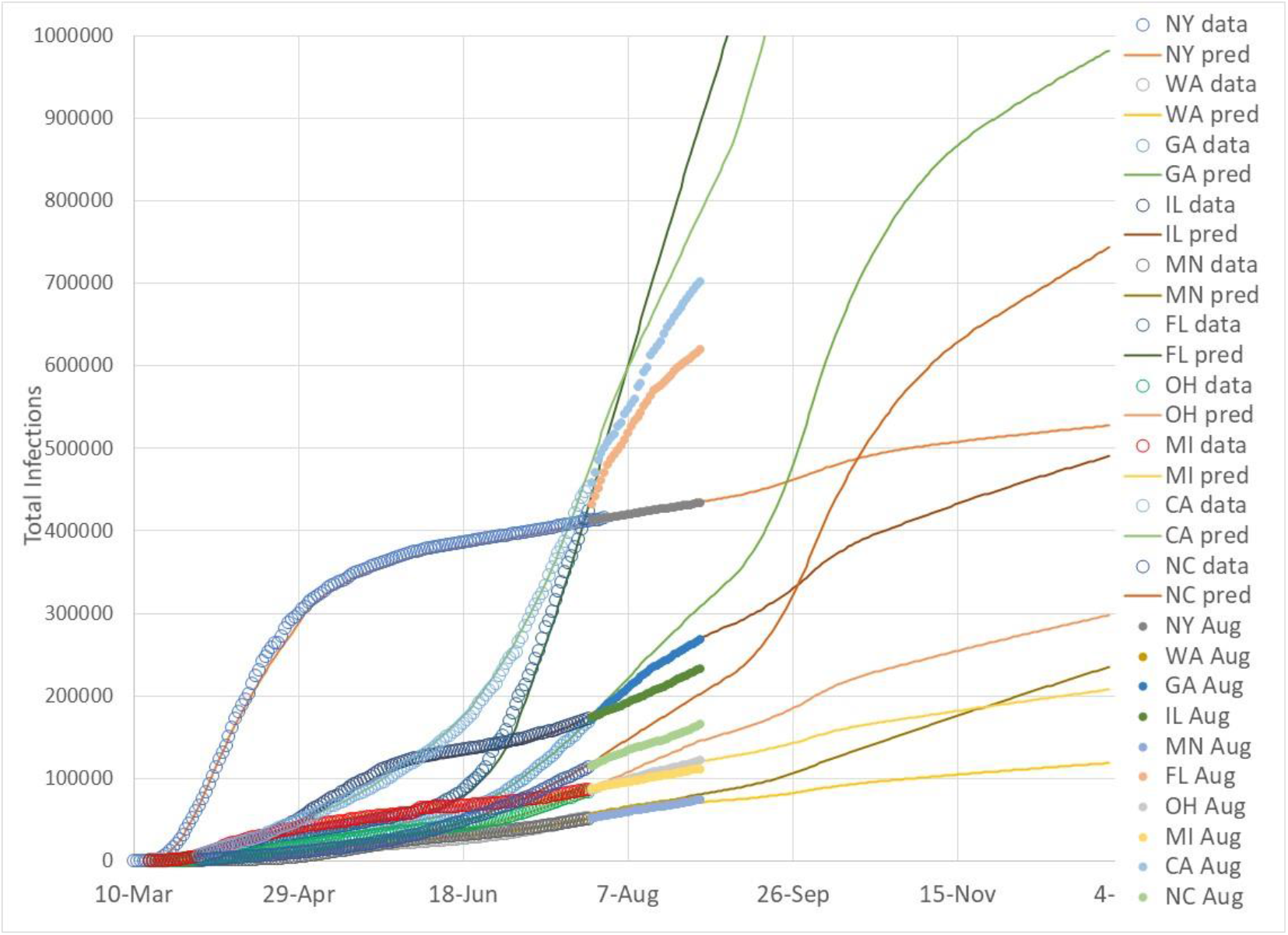
*Comparison of prediction and actual August total infection data for 10 State for School with Temperature effect*.

**Figure 9.**
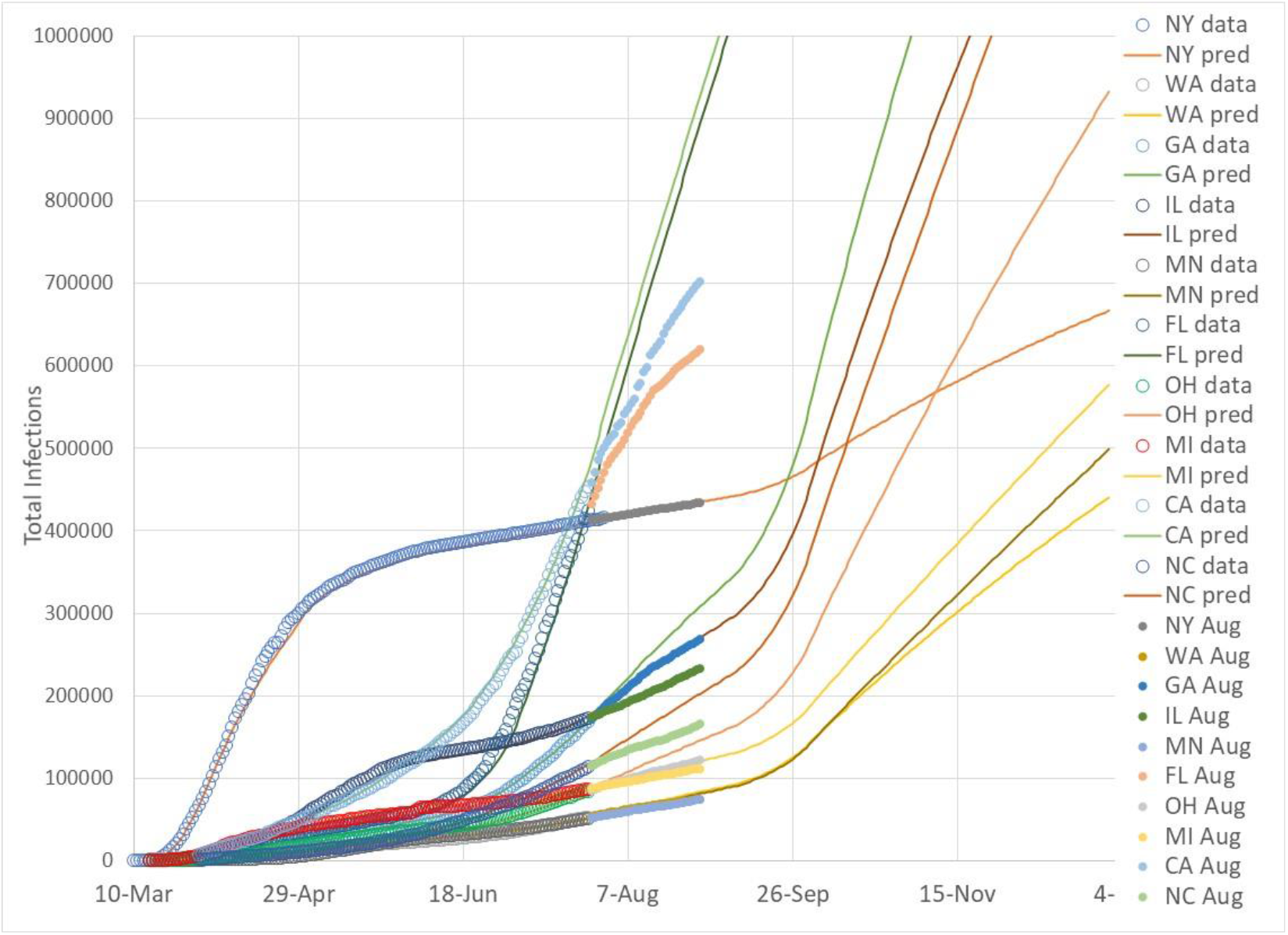
*Comparison of prediction and actual August total infection data for 10 State for School with No Temperature effect*.

Except for Washington and California, the other 8 States (NY, GA, IL, MN, FL, OH, MI, and NC) do not display significant differences among the four cases for August because physical school re-opening effects and outdoor temperature effects do not occur until September. Washington is already in the 50F (10F) to 70F (21C) temperature window, and removing the temperature effect has the impact of elevating predicted infections for Cases 2 and 4. California is assumed to have some temperature effect in August, and in a similar but reduced manner, displays an increase of infections when the beneficial temperature window effect is removed. Note that representing California as a single climate zone is not reasonable for those interested in forecasting infections for a specific region. The purpose of these predictions is to ascertain whether States that have taken very different infection pathways with more than an order of magnitude differences in infection cases can be represented by human behavior (SDI and G).

Case 1 most closely represents August by incorporating the outdoor temperature effect. Note that Case 3 is identical in predictions for August because the prediction model does not assume school re-opening effects until September, and therefore, Table values for Case 1 and Case 3 are identical, as well as Case 2 and 4 for all States except for WA and CA.

Table 1 prediction comparisons for New York and Washington are very close to actual infection cases as of August 31, 2020. NY and WA were the earliest and hardest impacted States during the emergence of Covid-19 cases in March 2020. Both States enacted strict control measures, and by the end of April had been able to sustain Infection Parameter levels below the 2.72 linear boundary, resulting in decreasing active infectious cases. Both NY and WA have gravitated toward 2.72 IP levels, however, both States have maintained vigilance and as the summer surge emerged in June, both States responded quickly and avoided the large increase in active infections. The close agreement of NY and WA predicted and actual infections at the end of August reflects both States moving along the linear infection growth boundary.

Case 1 (and Case 3) for the other 8 States show a systematic decrease of actual infection cases relative to the predict infections that is related to infection surge levels during June and July. Florida, for example, was severely impacted by the infection surge. In response to horrific news reports of rapid infection growth and hospital staffing shortages, strict control measures (eg, mandated face mask wearing) and greater adherence of the State’s populace to control measures moved Florida to sustained IP levels below 2.72, resulting in reduction of active infection cases that continues through August. The difference in predicted infections (920,599) versus actual infections (623,471) reflects the effectiveness of disease transmission control actions taken by Florida occupants during the month of August. Florida now appears to be moving upward in IP toward the linear growth boundary.

The remaining 7 States, to varying degrees (Michigan with 8% to North Carolina with 24% fewer actual infections than predicted) reflect the severity of the summer surge coupled with the State populace reaction to the surge severity. That is, lower summer surges required less severe control reactions, which resulted in those States not reducing IP levels significantly below 2.72.

Figures 2-4 demonstrate that the primary observation used to guide infection predictions continues to hold. All States (and countries) without strict mandated control measures gravitate toward a push-pull path that follows the linear IP boundary value of 2.72. And IP value is a function of two human behavior parameters; gross human interaction as characterized by the Social Distance Index (SDI) and inter-personal interactions as characterized by the disease transmission efficiency (G).

## Comparison of Actual Infection Data with Adjusted August Infection Predictions

The prediction model is adjusted for August for all States except New York and Washington. Adjustment is made to the disease transmission efficiency parameter during August for the 8 States. Beyond August, the prediction model’s parameters are unchanged through December 31, 2020. In this manner, month-by-month variations between prediction cases (school and outdoor temperature effects) can be examined.

Figures 10-13 show predicted total infections through December 31 with the August adjustments relative to actual infection data through July 31 (initiation of prediction model) and August 2020 actual infection data. Note that the same adjustment to the disease transmission efficiency parameter (G) was made for all four cases, and as previously described, the differences among the four cases are not significant except for WA and CA.

**Figure 10.**
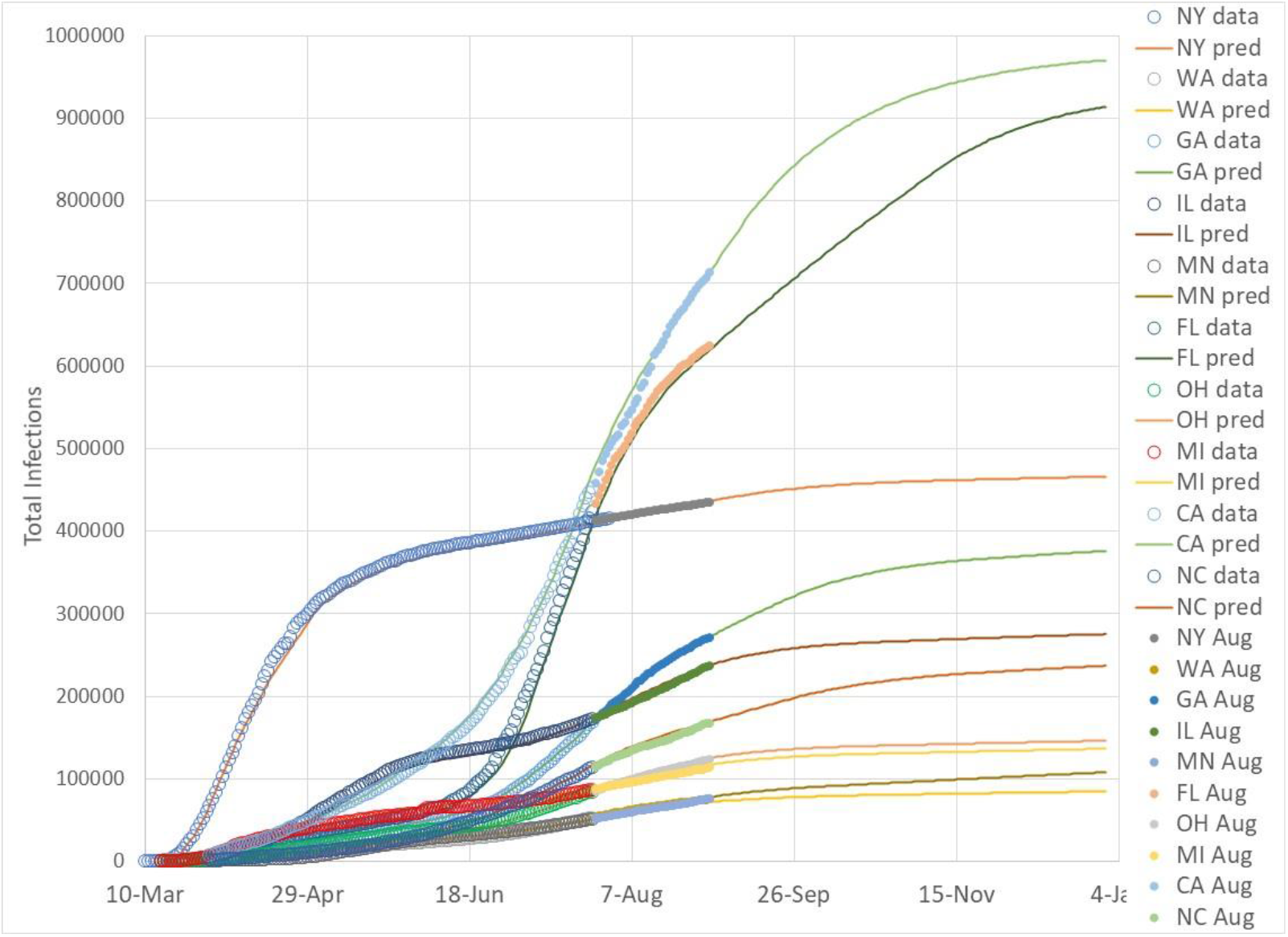
*Comparison of adjusted August2020 prediction and actual August total infection data for 10 State for No School with Temperature effect*.

**Figure 11.**
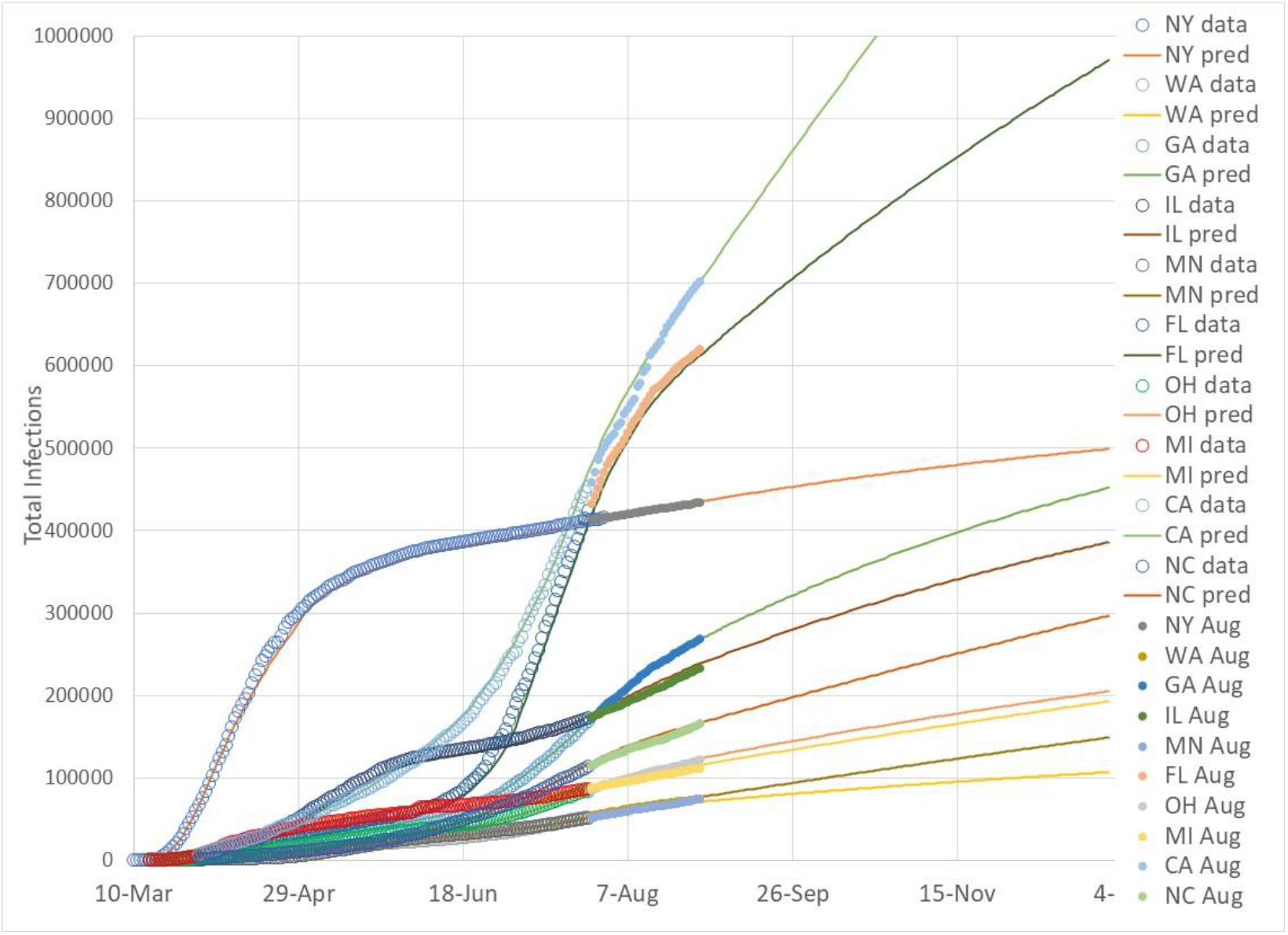
*Comparison of adjusted August 2020 prediction and actual August total infection data for 10 State for No School with No Temperature effect*.

**Figure 12.**
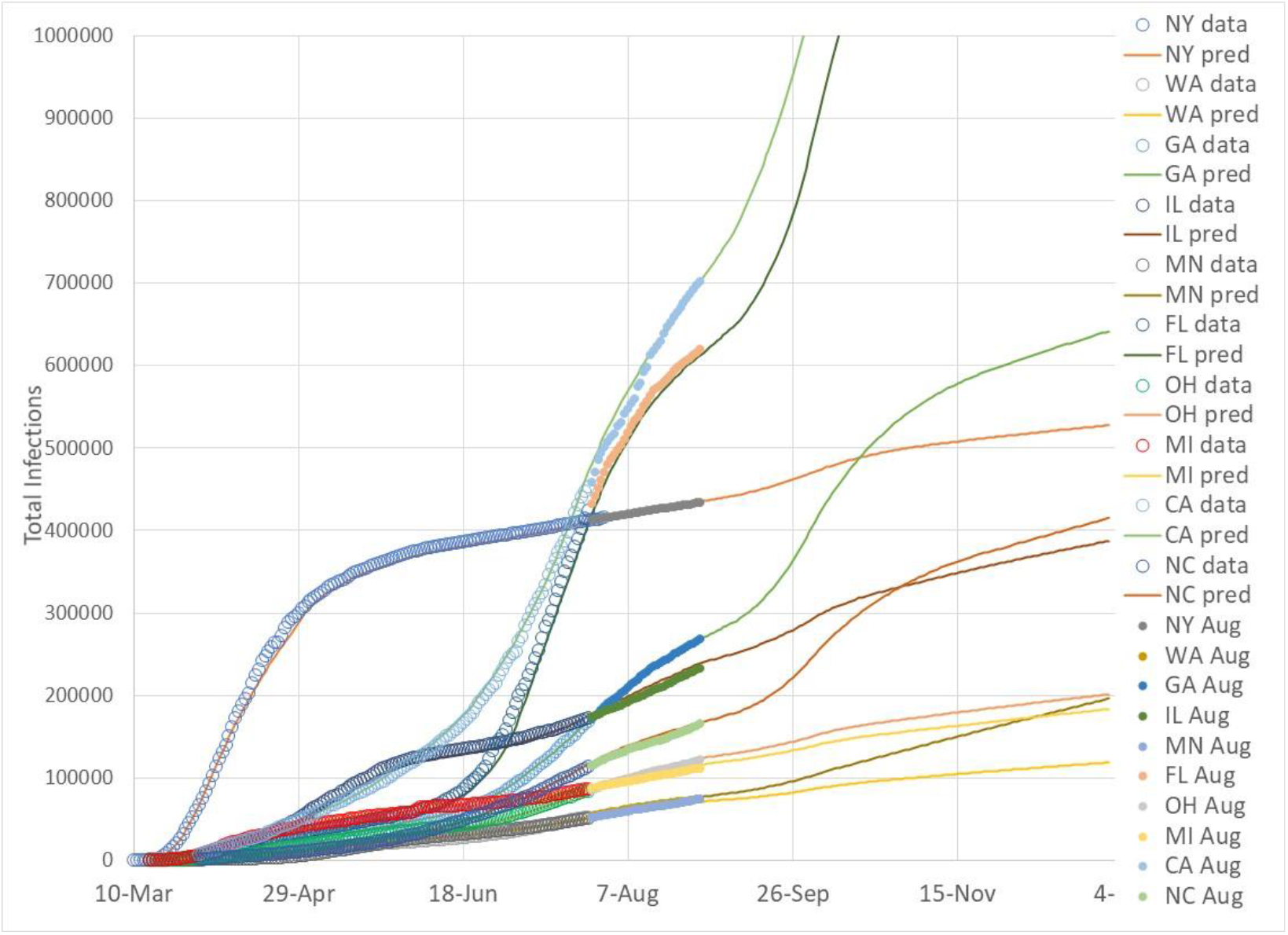
*Comparison of adjusted August 2020 prediction and actual August total infection data for 10 State for School with Temperature effect*.

**Figure 13.**
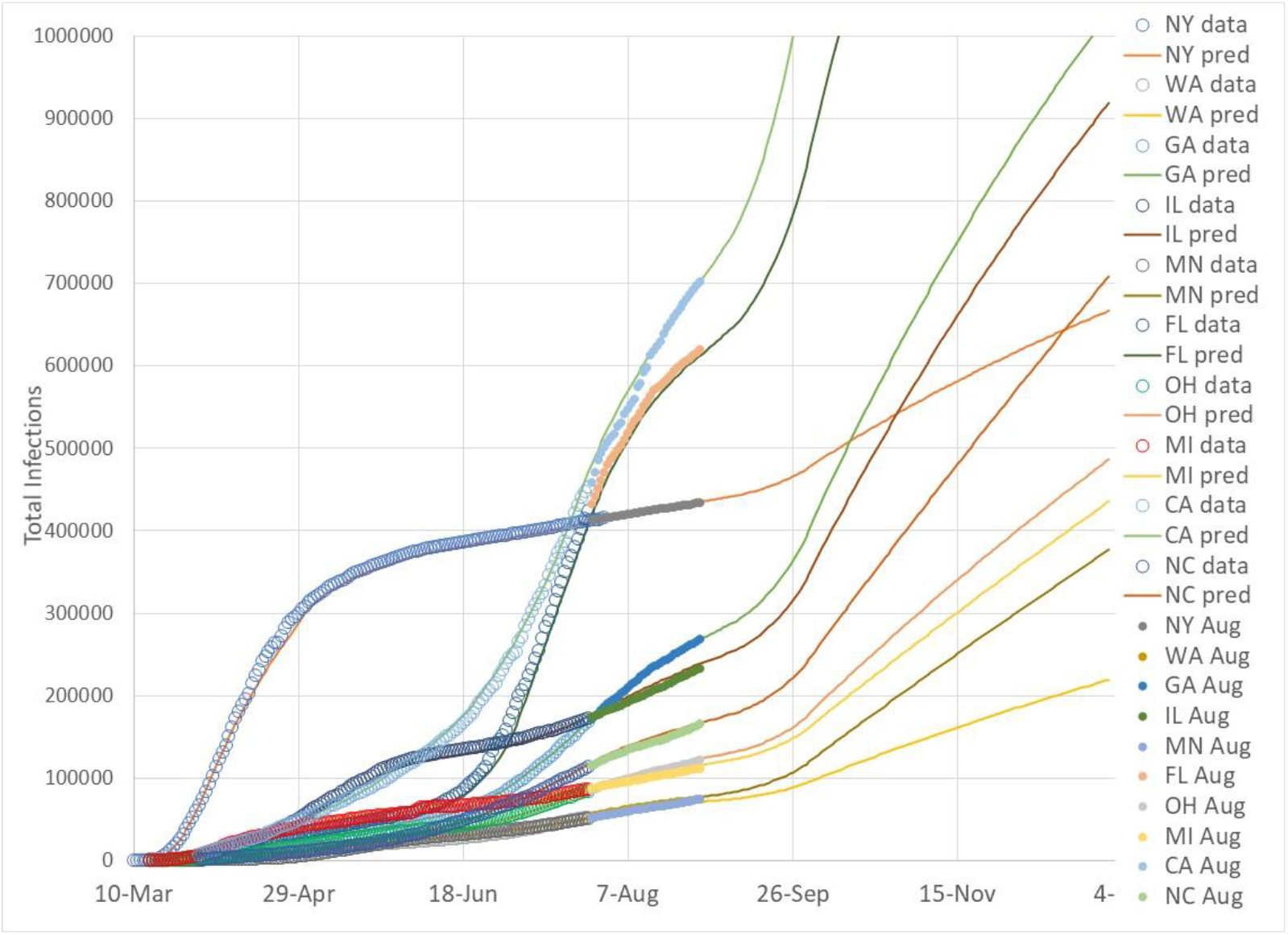
*Comparison of adjusted August 2020 prediction and actual August total infection data for 10 State for School with No Temperature effect*.

The importance of infection control measures undertaken in California and Florida during August are very significant and impact the trajectory of infection growth through the rest of the year. Both Florida and California would have exceeded 10,000,000 total infections by the end of September for all four cases if the States had not taken measures to reduce IP below 2.72 through August, reducing the current number of active infectious cases. Because of the actions taken, both California and Florida have the potential to keep total infections below 10,000,000 by the end of the year (Case 1, no school effect with temperature effect, Figure 10). Similarly, all other States with the exception of New York and Washington that have not been adjusted for August, reduced predicted infections that help reduce future infections through the rest of the year assuming an IP kept near the 2.72 linear boundary value.

## New Daily Infection Case Comparisons

New daily infection cases are important for understanding upcoming medical staffing levels. Daily infection cases are determined from the prediction model by subtracting a previous day’s total infections from the current day’s total infection prediction. Both real data and predicted data for new daily infections are “noisy” as expected when taking differences. Weekend versus weekday infections caused by daily personal interaction variations and other factors such as day-to-day data reporting differences affect new infection reports.

Figures 14-23 display predicted and actual new daily case data for each State. The predicted data is for Case 1 (No School effect with Temperature effect) using prediction data with August adjustments to the model (Figure 10 data). The figures are roughly ordered by magnitude of new daily infection cases. NY, FL and CA, shown in Figures 14, 15, and 16, experienced the highest new daily infection levels, and are scaled differently than the other 7 States. The simulation model begins in March 2020 with IP data that follows actual IP trends through August. Beyond August through December 2020, IP values are set just below the linear boundary limit (2.72) by selecting a combination of SDI and G as described in the previous report (1).

**Figure 14.**
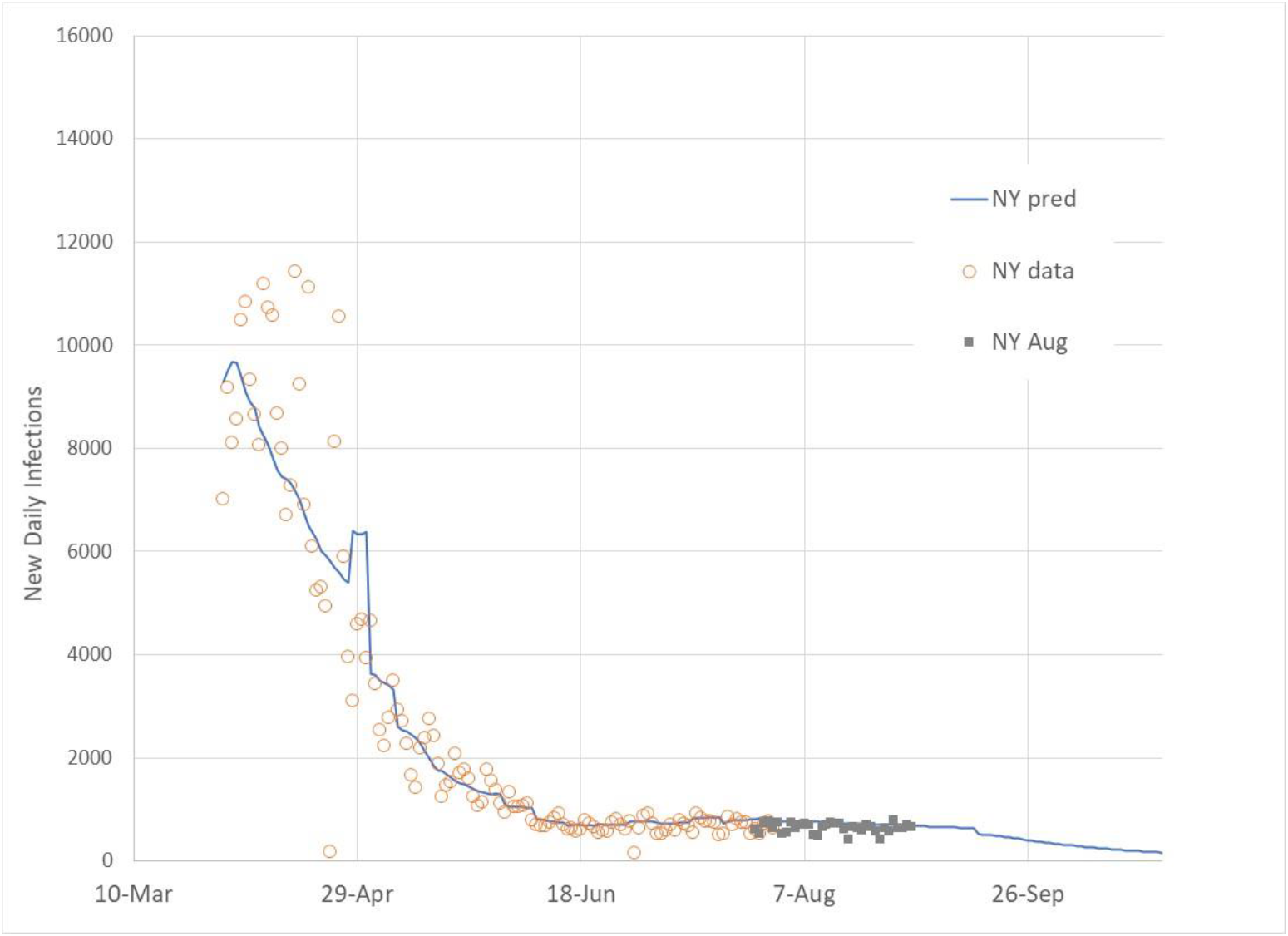
*New daily infection cases (actual through July 27, actual through Aug 31, and predicted) for NY*.

**Figure 15.**
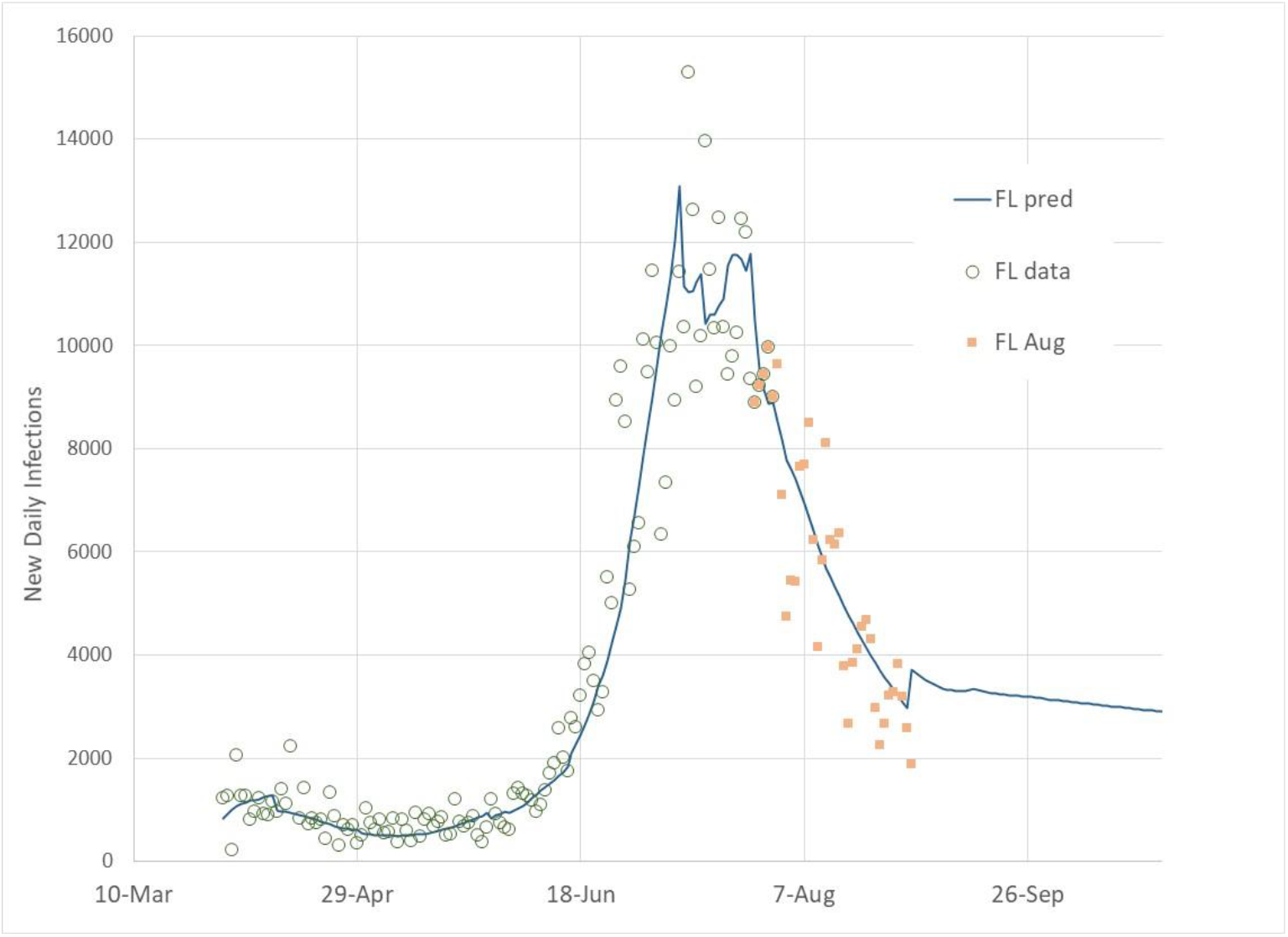
*New daily infection cases (actual through July 27, actual through Aug 31, and predicted) for FL*.

**Figure 16.**
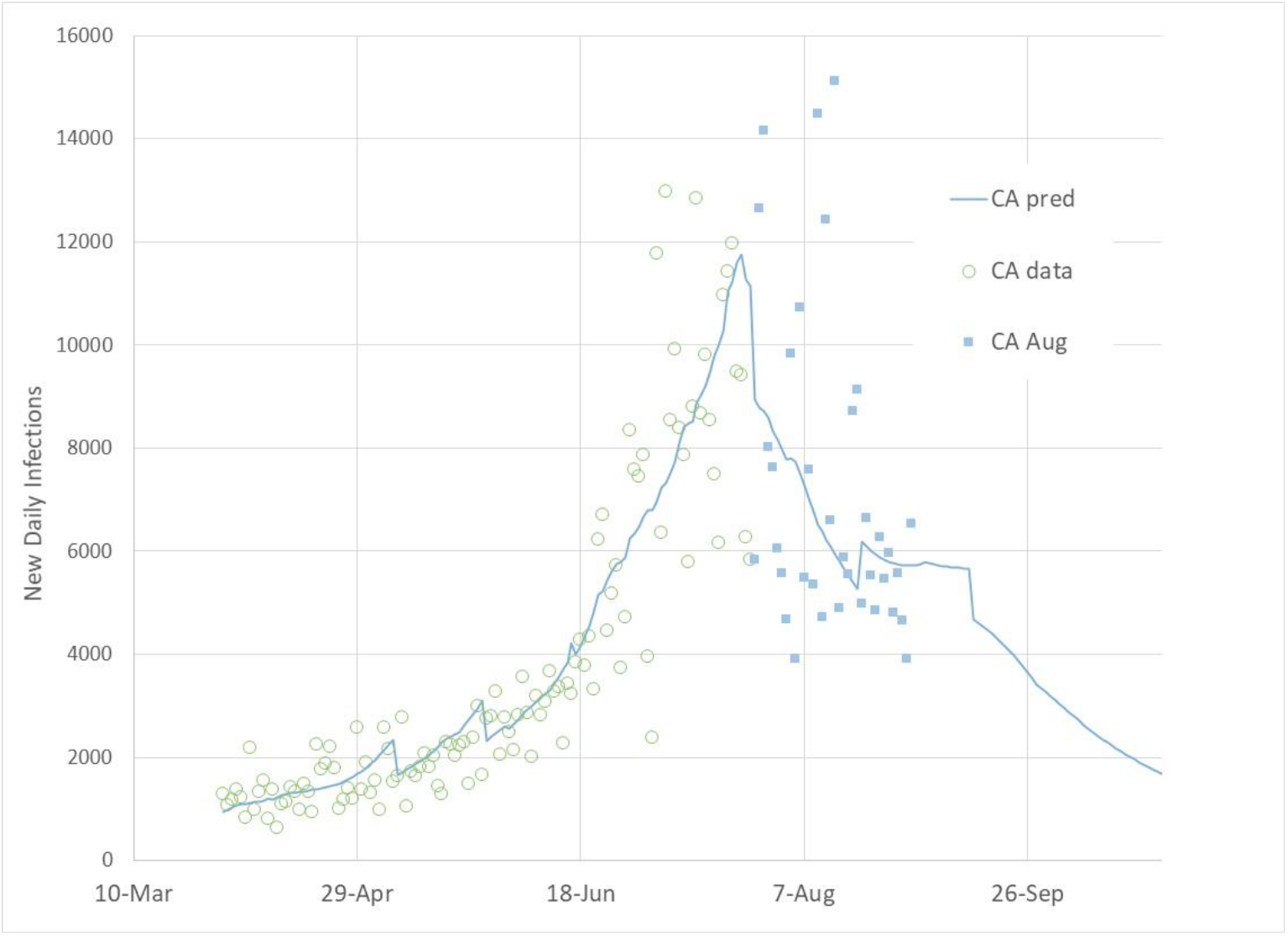
*New daily infection cases (actual through July 27, actual through Aug 31, and predicted) for CA*.

**Figure 17.**
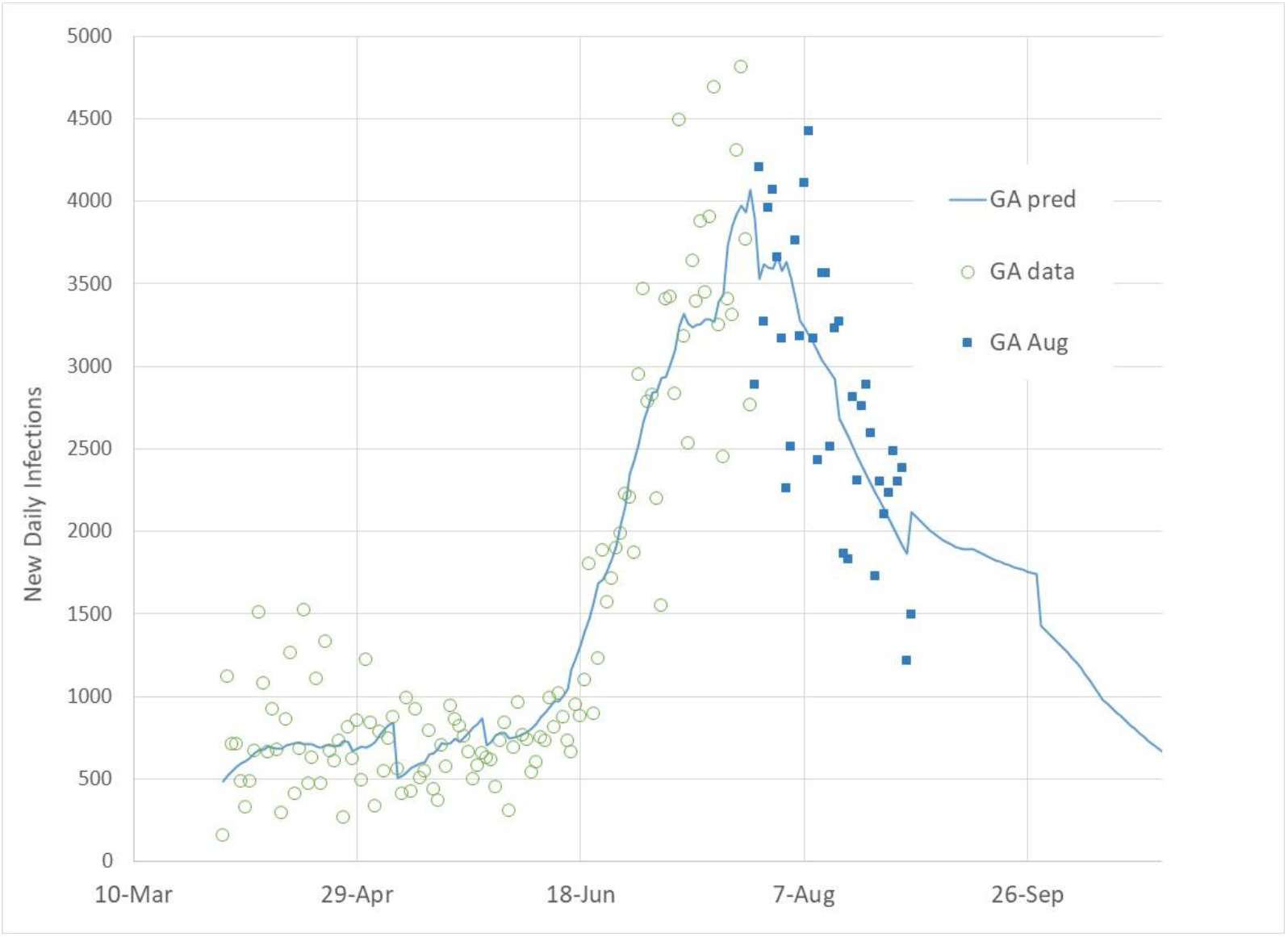
*New daily infection cases (actual through July 27, actual through Aug 31, and predicted) for GA*.

**Figure 18.**
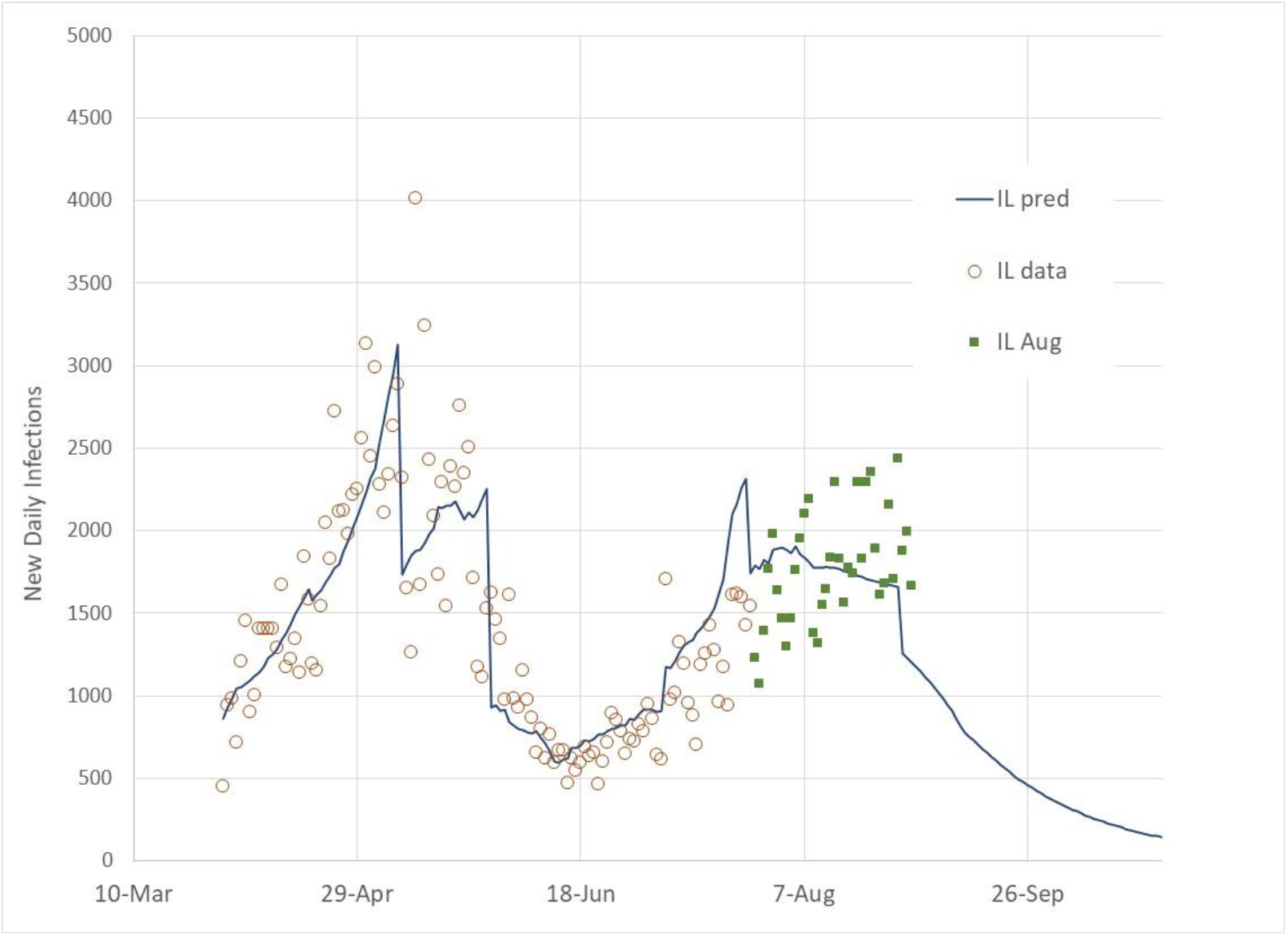
*New daily infection cases (actual through July 27, actual through Aug 31, and predicted) for IL*.

**Figure 19.**
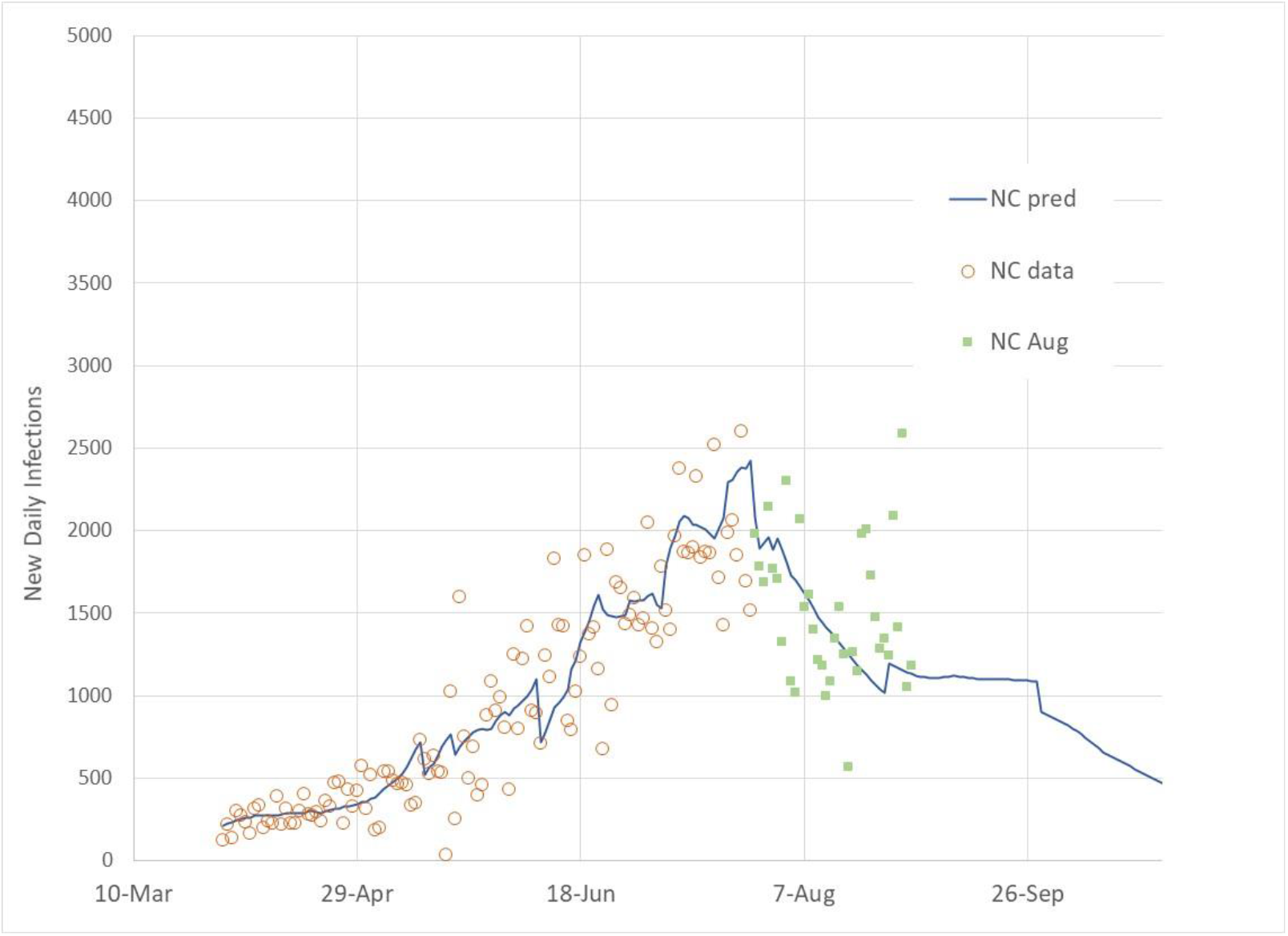
*New daily infection cases (actual through July 27, actual through Aug 31, and predicted) for NC*.

**Figure 20.**
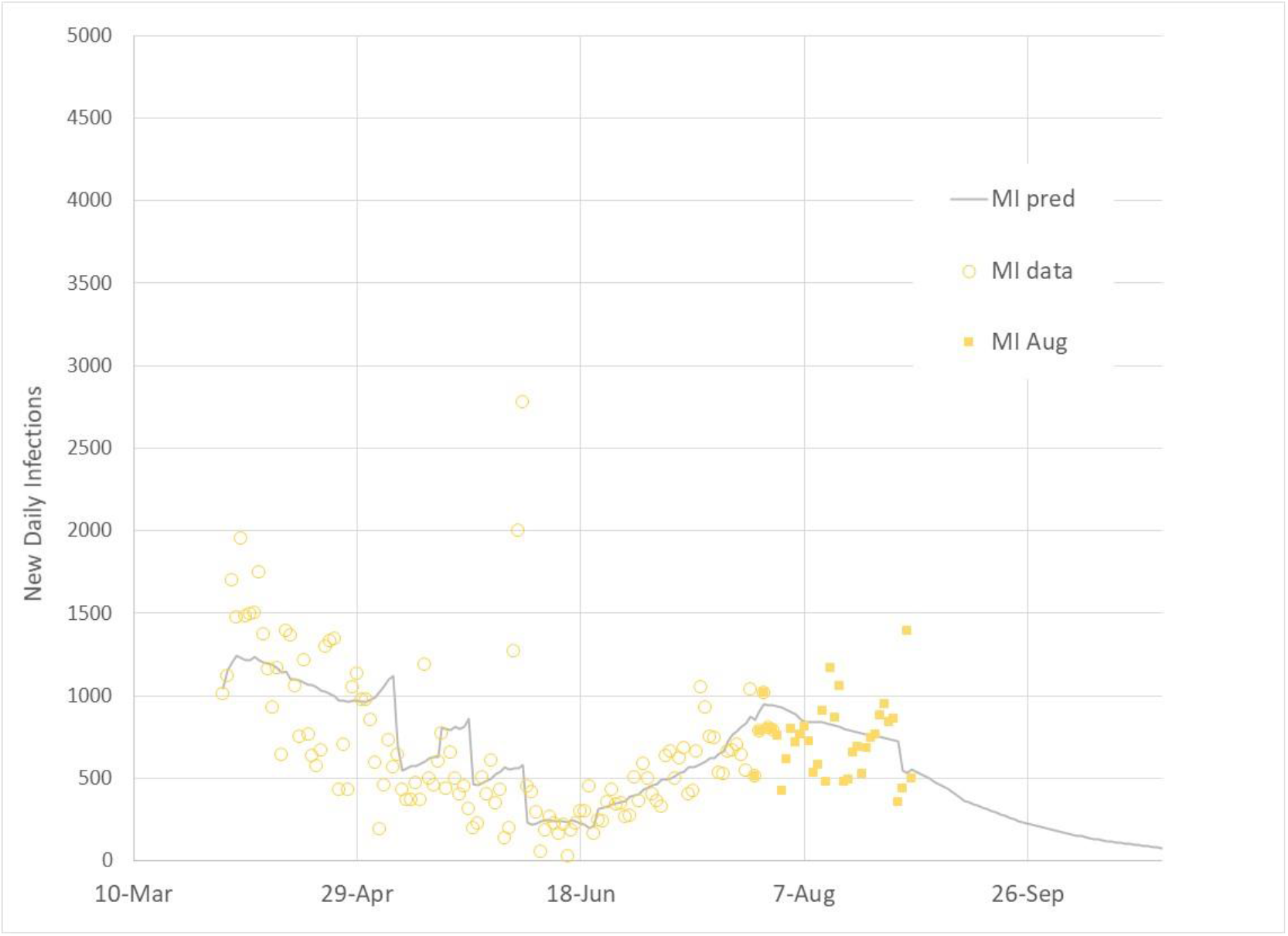
*New daily infection cases (actual through July 27, actual through Aug 31, and predicted) for MI*.

**Figure 21.**
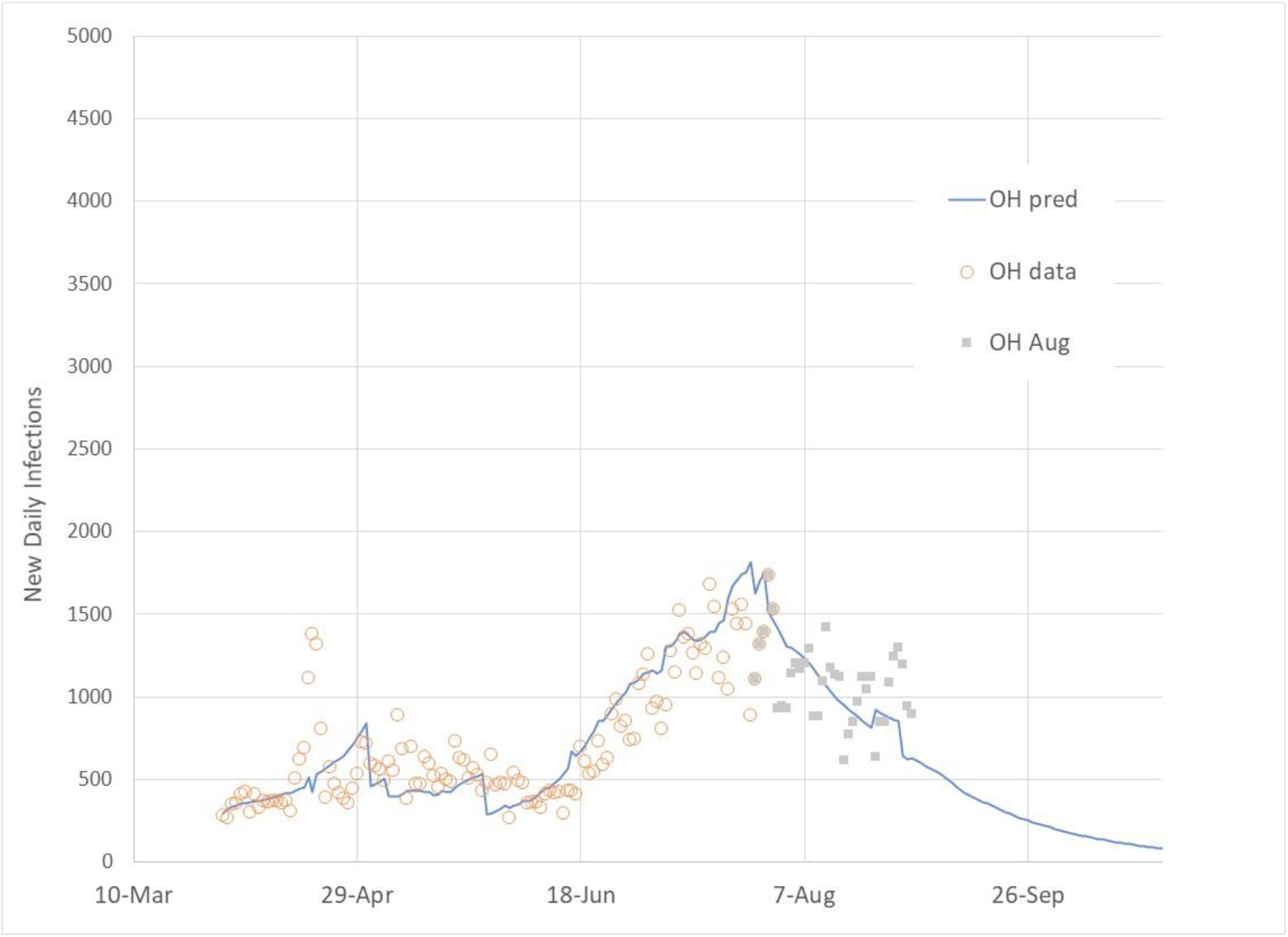
*New daily infection cases (actual through July 27, actual through Aug 31, and predicted) for OH*.

**Figure 22.**
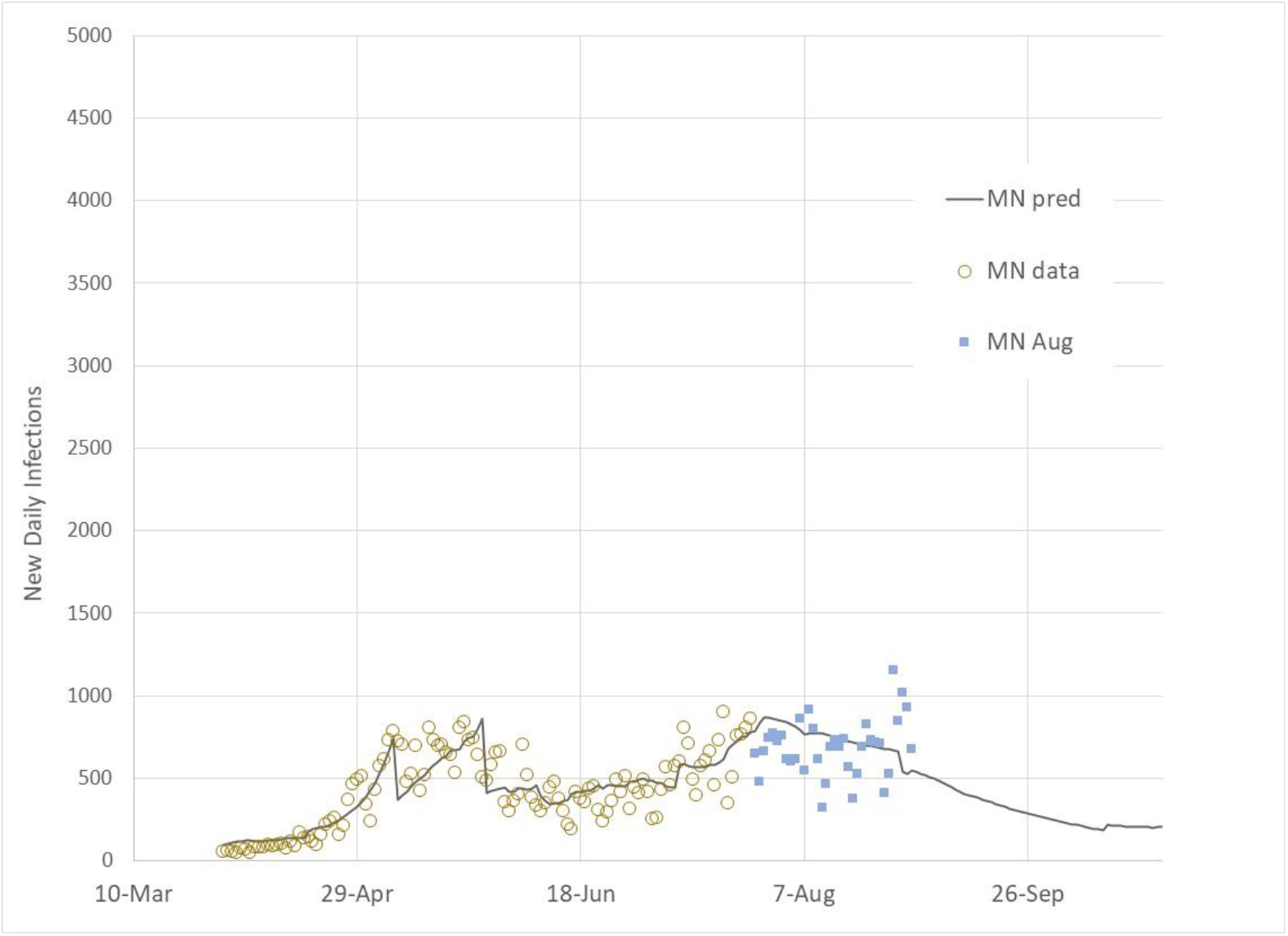
*New daily infection cases (actual through July 27, actual through Aug 31, and predicted) for MN*.

**Figure 23.**
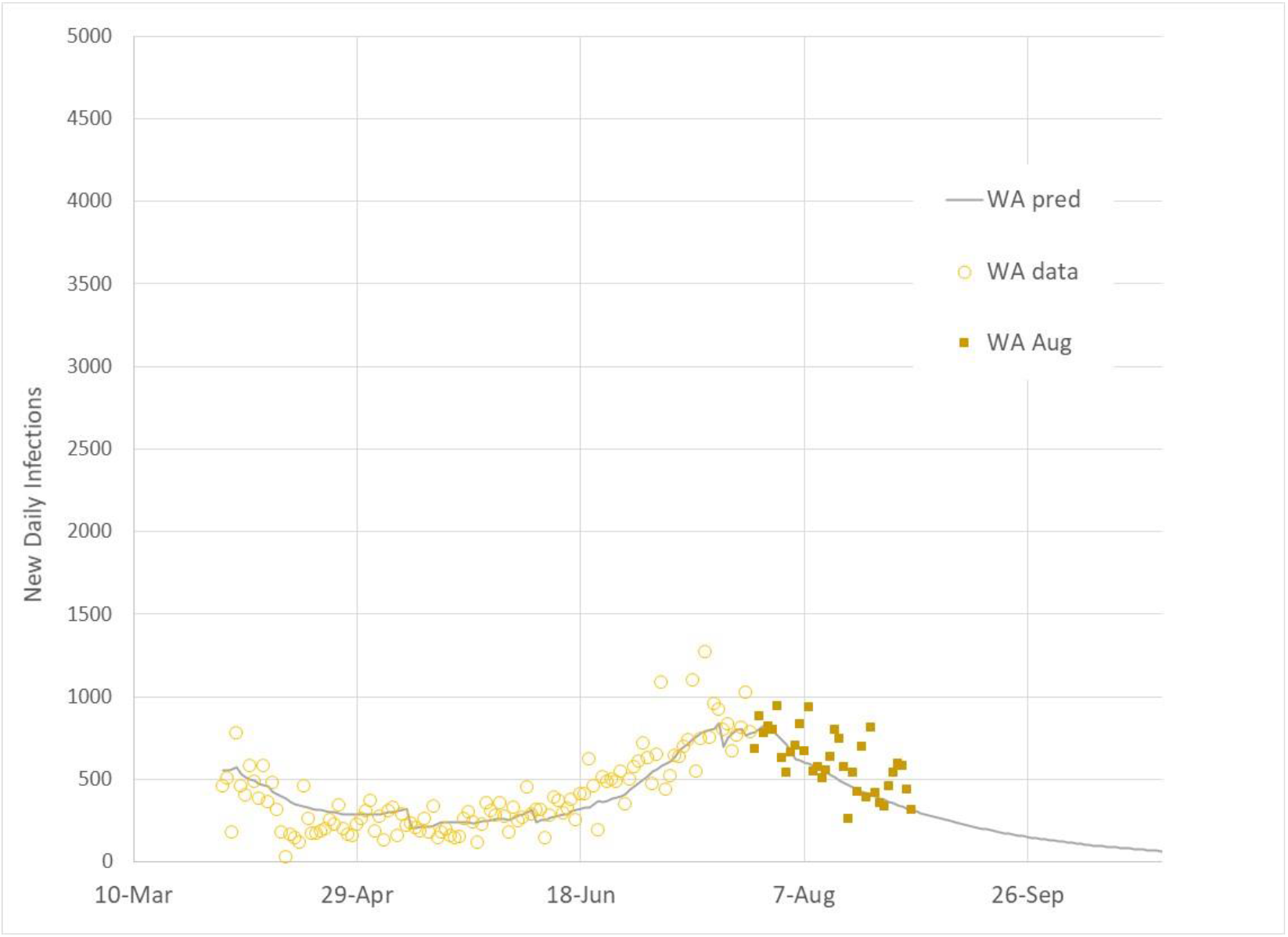
*New daily infection cases (actual through July 27, actual through Aug 31, and predicted) for WA*.

New York (Figure 14) experienced it highest new daily infections in March. Since April, New York has kept good control of the virus. Minimal new daily infection increases occurred in June and July unlike other States with strong summer infection surges. New daily infections continued at a steady level through August, reflecting New York’s movement along the linear IP boundary. Florida (Figure 15) and California (Figure 16) were impacted by the summer surge due to attempts to re-open businesses (decreased SDI) without strong interpersonal protection measures (reduced G). Both Florida and California continue to recover from the summer surge. Both States new daily infection cases continue to decrease, however, the rate of decrease is slowing indicating a movement toward the linear infection growth boundary.

Georgia’s (Figure 17) new daily infection path is similar to Florida and California with a strong summer surge, followed by decreasing new daily infections as more stringent infection controls were implemented. Georgia continues to decrease its new daily infection cases.

Illinois’s (Figure 18) new daily infection path is one of the most complex with high infection loading during the early emergence of the pandemic in March, and a strong regrowth of new daily infections during the June and July summer surge.

North Carolina’s (Figure 19) new daily infection path is similar to infection patterns in other southern states with poorly regulated business re-openings. Michigan (Figure 20), Ohio (Figure 21) and Minnesota (Figure 22) new daily infection trends are similar to Illinois in which outdoor temperature effects were the primary cause of their summer infection surges (2). Finally, Washington (Figure 23) experience a significant summer infection surge relative to their pre-June infection levels.

## Summary

August has been a month of several States gaining control of the mid-June through July summer surge. As of the end of August, States are moving toward the linear infection growth path that has been observed to be the natural path for countries and regions that oscillate from accelerating to decaying infection regions.

The adjustment of model prediction parameters to reflect August infection data re-initializes model predictions through the end of the year. No adjustments are made to prediction model parameters for September through December, and the basic premise that the US and its States will follow the linear infection boundary defined by an Infection Parameter value of 2.72.

September prediction comparisons for the four case combinations of outdoor temperature effect and school re-openings should provide insight into the strength of both factors on impacting the course of infections through the end of the year. West coast fires may add another factor of unknown impact. For example, highly polluted outdoor environments may limit social interactions which should decrease infection transmission. Infections may increase, however, as more people spend time indoors with inadequate ventilation and closer inter-personal interactions.

## Data Availability

All data sources are cited and are publicly available.

https://www.worldometers.info/coronavirus/

http://91-divoc.com/

https://data.covid.umd.edu

## Notes

### Competing Interest Statement

The authors have declared no competing interest.

### Funding Statement

No research support has been received for this work.

### Author Declarations

No IRB oversight is required for this work.

